# Inexpensive and colorimetric RNA detection by *E. coli* cell-free protein synthesis platform at room temperature

**DOI:** 10.1101/2021.11.29.21267025

**Authors:** Michela Notarangelo, Alessandro Quattrone, Massimo Pizzato, Sheref S. Mansy, Ö. Duhan Toparlak

## Abstract

We report colorimetric detection of SARS-CoV-2 viral RNA by an *in vitro* transcription/translation assay with crude *E. coli* extracts at room temperature, with the aid of body heat. Clinically-relevant concentrations of viral RNA (ca. 600 copies/test) were detected from synthetic RNA samples. The activation of cell-free gene expression was achieved by toehold-switch-mediated riboregulatory elements that are specific to viral RNA sequences. The colorimetric output was generated by the α-complementation of β-galactosidase ω-fragment (LacZω) with cell-free expressed LacZα, using an X-gal analogue as a substrate. The estimated cost of single reaction is <€1/test, which may facilitate diagnostic kit accessibility in developing countries.

## INTRODUCTION

Early days of COVID-19 pandemic has proved that rapid and efficient diagnostic tools are indispensable to cope with the emerging infections to mitigate the detrimental effects of lockdowns. To this end, many laboratories and companies around the world have deployed their resources towards generation of fast and cheap diagnostic tools^1–3^. In this context, cell-free protein expression (CFPS) platforms emerged as attractive tools, as they are relatively simple, cheap and straightforward to engineer^4–7^. Moreover, freeze-drying of cell-free components to rehydrate with aqueous samples enabled cell-free biomanufacturing as a possibility within reach^8–10^. Further efforts were diverted to adapt CFPS platforms for efficient virus-specific detection, in particular with CRISPR/Cas nucleases^11–14^. In parallel, multiple rapid, enzymatic RNA amplification methods were also developed with colorimetric output^15–17^. Many of these platforms detect viral RNAs with high specificity at attomolar concentrations; but also suffer from relatively high costs per run, since they rely on commercial reagents and need trained workers. Furthermore, they also require re-cycling or incubation at above room temperature for optimal efficiency, rendering them problematic for field applications.

During unexpected global pandemics, resources may become scarce and limited. If massive diagnostic scale-up is necessary for screening the entire population, relying on commercial sources of reagents can crucially limit the diagnostic capacity. As the emerging SARS-CoV-2 variants continuously remind us, fighting a highly infectious respiratory virus requires a global strategy. To facilitate this goal, we need accessible and inexpensive diagnostic tools that are easy-to-deploy and affordable, especially in developing countries. With these strategic goals in mind, here, we report the potential of detection of viral RNA sequences by repurposing the E. coli cell-free transcription/translation system with colorimetric output (Scheme 1). We adapted toehold-switch-mediated riboregulatory elements for gene expression activation, preceded by isothermal RNA amplification aided with body heat. Using minimal equipment and cell-free reactions operating at room temperature, high-attomolar (ca. 110 aM) concentrations of viral RNA were detected from synthetic samples. The colorimetric output was generated by α-complementation of β-galactosidase ω-fragment using an X-gal analogue as a color-changing substrate with enzyme activity. In principle, the colorimetric diagnostic platform can be coupled to magnetic-bead-driven RNA isolation, where a proof-of-principle was demonstrated from saliva. We estimate the total cost of the colorimetric detection assay to be ca. 0.72 euro/test.

**Scheme 1.**
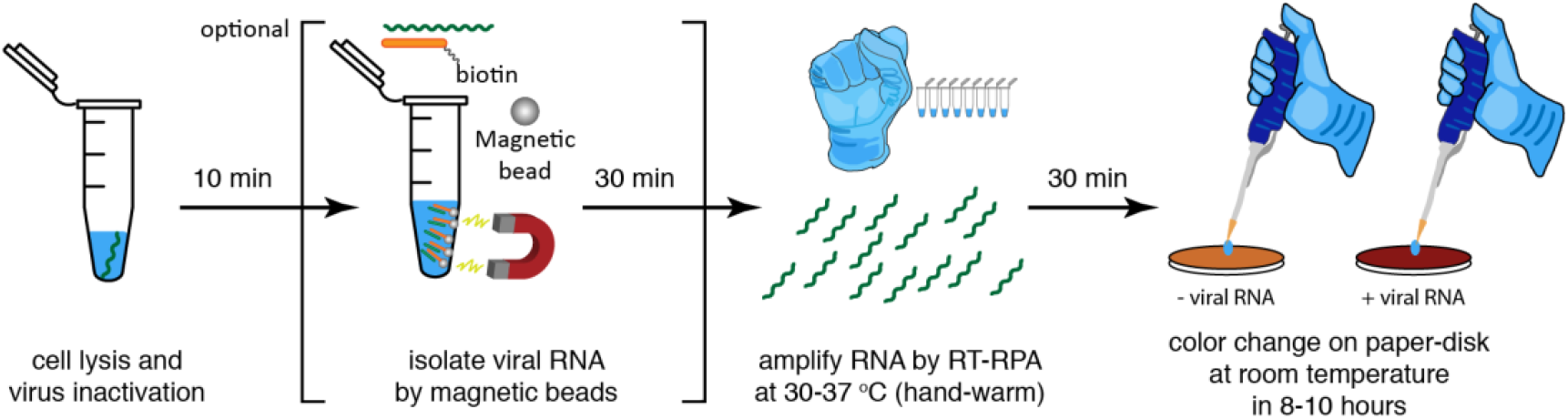
Conceptual summary of viral RNA detection by cell-free assays.

## RESULTS AND DISCUSSION

We *in silico* designed and functionally verified 11 different toehold-switch-mediated riboregulatory constructs that are complementary to the 5’ and 3’ untranslated regions (5’ UTR and 3’ UTR) of the SARS-CoV-2 genome (Figure S1). UTR was specifically chosen because, during the coronavirus replication cycle, discontinuous RNA synthesis generates higher-copy numbers of UTR sections than the rest of the genome (Figure S2)^18–20^. For initial characterization of the riboregulatory elements and cell-free extracts, we used superfolder GFP (sfGFP), controlled by T7 promoter (Figure 1A). The activation of cell-free gene expression with different toehold-switches was triggered by a complementary DNA oligonucleotide for initial screening. All riboregulatory elements activated the cell-free gene expression only in the presence of complementary oligonucleotides, but worked at different efficiencies (Figure 1B and S3A). The most promising construct, TH001, gave ca. 15-fold increase in gene expression, thus it was chosen for subsequent experiments. In *E. coli* cell-free systems, the innate nucleic acid detection limit, that is without any pre-amplification step, was found to be ca. 1 nM (Figure 1C and S3B). Such levels of toehold-switch activation are on par with the detection limit of PURExpress-based cell-free systems^6,21^. Thus, we concluded that a pre-amplification step was necessary to detect clinically-relevant concentrations of viral RNA.

**Figure 1.**
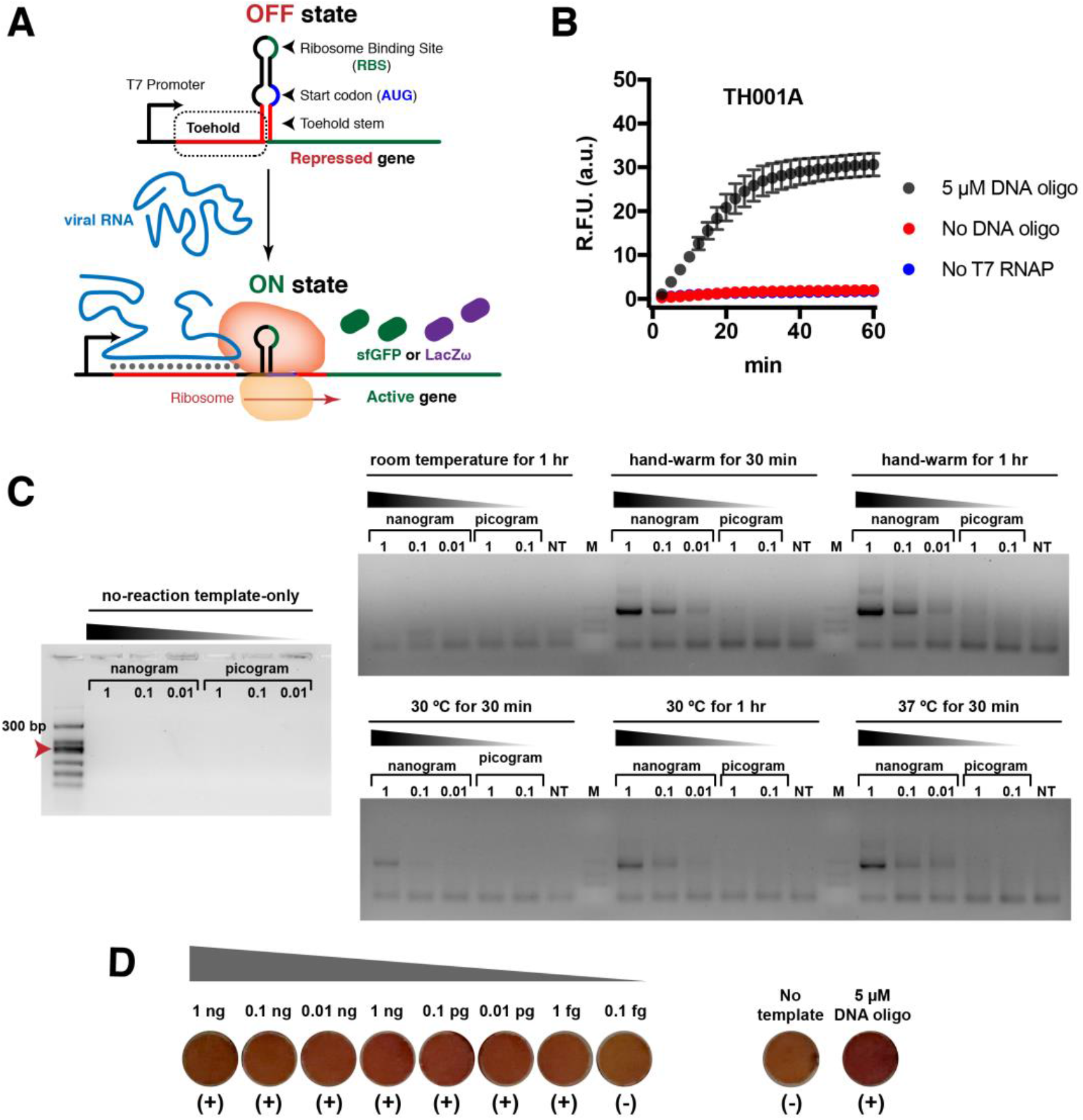
Colorimetric and cell-free RNA detection. **(A)** Toehold-switch-mediated activation of cell-free gene expression. **(B)** Activation of gene expression by the target viral sequences, provided as a DNA oligonucleotide. **(C)** Isothermal amplification of SARS-CoV-2 viral RNA at various conditions. Note that body-heat warmed (hand-held) reactions worked as good as 37 °C incubation. **(D)** Colorimetric assay reactions on-paper-disk from serial dilution of synthetic RNA with RT-RPA. NT ‘No Template’ stands for full cell-free reaction assembly but without any complementary nucleic acid present template reaction.

Point-of-care (POC) and inexpensive diagnostic tools may need exclusively room temperature operations, especially if instrumentation is scarce. To this end, cell-free reaction conditions and extract preparation protocols were tested for room temperature work-up (Figure S4). That is, *E. coli* extracts were prepared following post-log-phase growth at 23 °C. *E. coli* extracts were found to retain 50% activity compared to regular 37 °C growth (Figure S4A). To minimize the dependency to expensive instruments, the functionality of cell-free reactions was tested and verified without freeze-drying but only after drying (Figure S4B). This way, we potentially eliminated the need for expensive freeze-drying equipment for preparation of lyophilized cell-free reactions. At last, POC diagnostics may use one-step viral inactivation from bodily fluids, followed by the cell-free reactions. To this end, the cell-free reactions were tested for compatibility with human saliva. As a reaction additive, saliva did not inhibit the reactions, as long as dilution was used between 1/20^th^ and 1/100^th^ of the reaction volume (Figure S4C).

Following these optimization efforts of the cell-free extract, we focused on development of cell-free-compatible isothermal RNA amplification strategies. To this end, nucleic acid sequence-based amplification (NASBA)^22^, reverse transcriptase-recombinase polymerase amplification (RT-RPA)^23,24^ and reverse transcriptase rolling circle amplification (RT-RCA)^25^ reactions were tested (see supplementary materials). Out of all pre-amplification methods, RT-RPA proved to be the most efficient technique, both at recommended temperatures and at 30 °C (Figure 1C and S5). Nevertheless, we decided to put further effort in optimization of NASBA reactions, which can be assembled in-house and used in high-throughput sequencing-based diagnostics^26^. Our goal was to minimize the dependency of diagnostic tools to commercial components and kits –proven to be detrimentally scarce at the early days of COVID-19 pandemic.

We assembled NASBA reactions using past reports as guide^27,28^ and additionally tested different parameters. Our major goal was two-fold: (1) to couple NASBA to cell-free reactions; (2) to minimize temperature cycling and preferably operate solely at room temperature (22 ± 2 °C). Initially, we screened 10 different reaction additives inside a unique buffer composition that is suitable for all enzymes in the cocktail. (Figure S5A, supplementary methods). The additives that elicited the most positive impact were 10% (v/v) DMSO and 1 M betaine.

Past reports showed that “unoccupied” T7 bacteriophage RNA polymerase (T7 RNAP) can trigger transcription of random, non-specific RNA duplexes^29^. In order to alleviate this issue, random DNA oligonucleotide duplex was added to the mixture to minimize non-specific transcription by T7 RNAP (Figure S5B)^30^. The NASBA reaction conditions were further tested with decreasing enzyme concentrations at room temperature (Figure 5C). Yet, none of the methods allowed for efficient RNA amplification at room temperature, including RT-RPA (Figure S5C and S5D). However, conditions for optimal enzyme activity in pre-amplification steps were conductive to a putative single-pot lysis of human cells and extraction of viral particles. Coupling with lysis buffer, the presence of 0.5% (v/v) Triton X-100 in the NASBA reaction mixture improved the amplification efficiency (Figure S6A). Nevertheless, the process of cell lysis from human saliva did not overlap with RNA amplification in NASBA reaction, as a “ready-to-use” reagent (Figure S6B).

To minimize dependency of temperature cycling, we further tested NASBA without pre-heating at 65 °C for primer annealing. A pre-heating step proved to be non-detrimental for amplification, albeit at a cost of decreased efficiency (Figure S6C). Given the intrinsic toxicity of low concentrations of glycerol for *E. coli* cell-free expression system^31^, and well-known sensitivity of T7 RNAP to reducing storage conditions, we conclude that a system independent of T7 RNAP is the best choice for reproducible results. In the end, the lysis buffer composition was also more compatible with RT-RPA than NASBA (Figure S6C) to single-pot purification and amplification of viral RNA from saliva samples. Moreover, since field conditions require minimal instruments, we confirmed that RT-RPA works as good as 37 °C, when 8-strip tubes were hand-held and warmed by body heat (Scheme 1 and Figure 1C). We find this approach significant, since healthcare workers or the patients themselves can perform the test without any instrumentation apart from micro pipettors.

Having determined the most conductive pre-amplification mode as RT-RPA, we set out to modify our reporter system from fluorometric to colorimetric output. To this end, maltose-binding protein (MBP)-tagged β-galactosidase ω-fragment (LacZω) was expressed in *E. coli* NEB5α strain, purified via Amylose Resin and rescued from MBP by TEV protease digestion (Figure S7A). For α-complementation, LacZα subunit gene expression was placed under the control of endogenous *E. coli* promoters (Figure S7B and Table S1). To test the complemented LacZ activity, 2 mM (final or 12 µg/µL) Chlorophenol Red-β-D-galactopyranoside (CPRG) was used as a substrate to give an expected color change from yellow to reddish-purple (we note that the precise color change is heavily dependent on the solid support)^32^.

*E. coli* BL21-derivative strains are regularly employed for cell-free extract preparation and contain an endogenous copy of LacZ, which was not suitable for α-complementation (Figure S8A) To this end, we set out to prepare cell-free extracts from *E. coli* strains of JM109 and DH10β, which have genotypes with a LacZΔ15 mutation. The cell-free gene expression from the crude extracts of JM109 and DH10β was verified and, in the presence of toehold-switch triggering complementary DNA oligonucleotide, the colorimetric assays showed the color change from yellow/orange to orange/red within 1 hour, demonstrating the functionality of our reporter system (Figure S8C).

Having shown the cell-free reactions generated colorimetric output at room temperature, we next set out to couple RT-RPA amplification step to cell-free reactions. First, RT-RPA was run for 30 min at 37 °C and then added to cell-free reaction at a 1:20 dilution. Within 8 hours after rehydration, the reddish color was developed down to ca. 110 attomolar (aM) final concentration of RNA, that is ca. 667 copies of RNA per reaction, which was verified by qRT-PCR having cycle threshold (Ct) value higher than 28.79 ± 0.36 (Figure 1D and S9). In other words, as long as the RT-RPA amplified RNA generated a Ct value above 10-12, we detected a color change (Figure S10A).

Subsequently, we wanted to render all steps compatible at room temperature, in a proof-of-concept experiment: (1) cell lysis + viral inactivation, (2) RNA capture in saliva, (3) RT-RPA and (4) cell-free colorimetric assay. We chose saliva as a biofluid for the test, since saliva can be autonomously self-tested by the users in a non-invasive manner. Synthetic RNAs were spiked-in human saliva and isolated by magnetic bead separation. Nevertheless, the isolated RNA could not be efficiently amplified with RT-RPA, suggesting an incompatibility in buffer compositions, as the effect of saliva on RT-RPA was not completely inhibitory (Figure S6C). In the end, we examined the clinical relevance of the proposed magnetic-bead isolation. That is, we tested whether we can reveal the presence of viral RNA in patient samples. First, positive patient samples were identified by 5’ UTR-specific qRT-PCR (1 hit out of 7 samples) and later samples were amplified by RT-RPA (Figure S10B). The amplified RNA was isolated from saliva spiked-in samples, verifying our approach in-principle, but requiring further optimization efforts to combine all the steps together in one-pot. The clinical translation of our findings, in part, can be achieved if the subsequent research maintains the goal to reduce the duration of the cell-free reactions as well as the target reaction costs at low levels after scale-up.

## CONCLUSIONS

Here, we reported colorimetric detection of clinically relevant concentrations of RNA with a low-cost cell-free assay, with all operating conditions at room temperature. In theory, this assay does not require any instrumentation apart from the micro-pipettors (and magnetic racks). The current system may be suffering from suboptimal reaction conditions and *E. coli* extract compositions. Such focused efforts are likely to decrease the detection limit to low-attomolar concentrations and even reduce the incubation times. We estimate the overall cost of the single test as low as ∼0.23 euro (see Supplementary Text and Table S2), including the labor costs to perform the test. Given that future efforts can be diverted to optimization of this assay as an end-user-friendly diagnostic, further cost reductions can be anticipated. At the moment, largest limiting factor appears to be the commercial dependence to RT-RPA reactions. Nevertheless, low µL volume RT-RPA and future scale-up efforts for *E. coli* extracts can also bring the assay costs substantially lower than our estimates, up to 50% reduction^10^. Finally, we excitedly point out that the colorimetric detection can be coupled to cell phone applications^11^ or wearable devices^17^, bringing cell-free synthetic biology technologies in our everyday lives. The clinical and translational potential of paper-based biosensors has once more highlighted with our work, given the raised limitations were addressed and optimization studies were performed.

## METHODS

### *E. coli* Cell-free Extract Preparation

Homemade *E. coli* cell-free extracts that are used for in-solution reactions were prepared from Rosetta 2(DE3) Singles strain (Novagen), using published protocols as a guide^33^. For on-paper reactions, the *E. coli* strains JM109 and DH10β were used. This crude extract preparation did not include the dialysis step. For details, see supplementary materials.

### Isothermal Nucleic Acid Amplification

Reverse Transcriptase Recombinase Polymerase Amplification (RT-RPA) reactions were by from TwistAmp^®^ Basic kit, and assembled according to manufacturer’s instructions (TwistDx). The reactions were assembled in 10 µL volumes and contained 1 µL of template (either clinical samples or synthetic RNA) and 0.2 µL (40 U) of RevertAid Reverse Transcriptase (ThermoScientific). The final RT-RPA volume was 0.525 µL in a 10.5 µL cell-free reaction (1:20 dilution). For details on the NASBA and RT-RCA, see supplementary materials.

### In vitro Transcription/Translation

All *in vitro* transcription–translation reactions were performed in a final volume of 10.5 µL. For in-solution reactions, the sfGFP fluorescence was measured by Rotor–Gene Q qPCR machine (Qiagen) or by a multi-well plate reader (Varioskan, ThermoFisher). For on-paper reactions, the amino acid solution mix and energy solution compositions were taken from the literature^33^. The supplementary solution additionally contained 10 mM maltose, glutamate salts (Mg^2+^ and K^+^) and 2% (w/v) PEG4000 as a molecular crowding agent. Purified LacZω (50 ng/µL) and LacZ substrate CPRG (0.12 µg/µL). Template plasmid DNA was at a final concentration of 30 nM.

### General Procedure for Colorimetric Assay

Prior to on-paper reaction, 1 µg of MBP-LacZω was digested by 1 µL TEV protease (New England Biolabs) in a 50 µL at 30 °C for 5 h. Then, 5% (w/v) BSA-blocked, air-dried (16 h) paper-disks were put in in clear bottom 96–well plate (Costar) and the *E. coli* cell-free reactions were added on top. Both synthetic RNA (pre-amplified) and clinical samples (purified and pre-amplified) were provided at a 1:20 final dilution. Then, the reactions were let air-dry and monitored for color change at room temperature.

### Synthetic and Clinical RNA Sample Preparation

The sequence of 5’ UTR(+) region of the wild-type SARS-CoV-2 was taken from NCBI (265 bp), obtained as a dsDNA gene fragment (GenScript) and cloned into an expression cassette under the control of consensus T7 promoter. *In vitro* transcription was performed by T7 RNA polymerase for 12 h at 37 °C from PCR-amplified template. Synthesized RNAs were initially cleaned up by TRIzol, ethanol precipitated and subsequently purified by NucleoSpin RNA Clean-up kit (Macherey-Nagel).

The RNA from infected patients was derived from anonymized saliva samples collected with ethical committee approval of the Azienda Provinciale per i Servizi Sanitari of the Autonomous Province of Trento (P.A.T.). Prior to the proof-of-concept RNA spike-in tests, saliva was diluted up to 5 or 10 times with PBS to reduce its viscosity.

Magnetic-bead-mediated RNA isolation was by biotinylated complementary DNA oligonucleotides to capture, and streptavidin-coated magnetic beads to isolate with a magnetic rack (Invitrogen). The experiments were performed using manufacturer’s instructions as a general guide, i.e., high-salt buffer to bind and low-salt buffer to wash; with significant modifications as specified in supplementary methods.

### Real-Time Quantitative Reverse Transcriptase PCR (qRT-PCR)

Prior to qRT-PCR, cDNAs were synthesized by reverse transcription with iScript™ cDNA synthesis kit (Bio-Rad). For clinical samples, the template was 14 µL (max. amount in 20 µL). qRT-PCR was performed by SsoAdvanced™ SYBR^®^ Supermix (BioRad). qPCR was run at Bio-Rad CFX96 Real-Time machine and acquisition was at FAM channel. qRT-PCR primer pairs were designed by online software Primer3, optimized for T_m_ = 57–60 °C, to generate an amplicon size of ca. 100 bp. Standard curve was generated by serial dilutions with 1:10 and primer efficiency was calculated as the slope of cycle threshold vs dilution factor.

### Genetic Constructs and Protein Purification

The toehold switches were designed using previously published principles (details in the supplementary methods)^34^. All clonings were performed by homemade Gibson Assembly mix^35^. All expression plasmids were with pSB1A3 backbone from iGEM Parts Registry. dsDNAs were obtained by PCR, gBlocks or in-house assembly of DNA primer-stitched templates. LacZω was cloned into a modified pMAL-c4X backbone containing N-terminus Maltose Binding Protein (MBP) with TEV protease recognition site flanked by (GS)_2_ linker sequence. *E. coli* NEB5α strain transformed with MBP-TEV-LacZω expressing plasmid was grown in Terrific Broth at 37 °C. The fusion protein was overexpressed with Autoinduction Medium^36^ with overall growth of 24h. The MBP-fusion protein was purified by Amylose Resin (New England Biolabs), eluted with 10 mM maltose.

## Data Availability

All data produced in the present study are available upon reasonable request to the authors.

## ASSOCIATED CONTENT

The Supporting Information is available free of charge on the ACS Publications website (link). The PDF contains details on methods, supplementary text, figures and tables.

## AUTHOR INFORMATION

### Author Contributions

Ö.D.T. conceived the idea, designed the study and wrote the manuscript. Ö.D.T. and M.N. performed the experiments. Ö.D.T., M.P., S.S.M. and A.Q. supervised the study. Ö.D.T. and S.S.M. acquired funding. All authors contributed to writing of the manuscript and given approval to the final version.

### Notes

No competing financial interests have been declared.

## ACKNOWLEDGMENT

Authors are grateful for Serge Nader, Anna Helander and Mirko De Pascalis for logistic support. We thank Max Mundt and the members of Mansy/Quattrone Labs for their comments on this work.

## Supplementary Materials for

### Supplementary Methods

#### Materials General

All *in vitro* transcription/translation components were purchased from Sigma–Aldrich (Merck) with highest possible purity, unless otherwise noted. The β-galactosidase (LacZ) substrate Chlorophenol Red-β-D-galactopyranoside was from Cayman Chemicals. *E. coli* Rosetta 2(DE3) Singles strain (Novagen) was used for the preparation of the generic *E. coli* extract for in-solution reactions. *E. coli* NEB5α (New England Biolabs) was used for general molecular cloning and recombinant protein expression. *E. coli* JM109 (Promega) was used for on-paper reactions. Whatman® α-cellulose filter papers were from Sigma-Aldrich (Merck) and cut with a generic paper hole puncher.

#### *E. coli* Cell-free Extract Preparation

Homemade *E. coli* cell-free extracts were prepared using published protocols as a guide^33^. Strain-of-choice was cultured overnight (15 h) in 2xYT+P without antibiotics, except for Rosetta 2(DE3) Singles strain (with chloramphenicol 34 µg/mL), initiated from freezer stocks^37^. The next day, the culture is transferred to larger volume conical flask with pre-warmed growth media with 1:100 dilution and incubated with shaking (220 rpm) without disturbing for 3.5 h (temperature varies). The cells were then harvested by centrifugation for 10 min at 6000 g, 4 °C. The bacterial pellets were briefly washed, resuspended in pre-chilled S12A buffer (14 mM Mg^2+^–glutamate, 60 mM K^+^–glutamate, 2 mM DTT, 50 mM Tris–Cl, pH 7.7 adjusted with concentrated acetic acid), centrifuged at 6000 g, 4 °C for 10 min. Following, the residual buffer was removed after another round of centrifugation at 6000 g, 4 °C for 2 min, and the wet pellet weight was determined. S12A buffer (0.9x dry cell weight) and 100 µm diameter glass beads were added and mixed thoroughly (5x dry cell weight). The resulting slurry–bead mixture was carefully transferred to bead beating tubes using 1 mL syringes without a needle. Bead beating was performed thrice for cell lysis, at a beat rate of 6.5 m/s for 30 s in cold-room (MP Biomedicals). The extract was separated from the glass beads by centrifugation at 6000 g, 4 °C for 10 min, with Bio–Rad Bio–Spin columns. The yellow-colored crude extract was then transferred to 2 mL tubes to remove the cellular debris by centrifuging once at 12000 g 4 °C for 10 min. The extract was then incubated at 37 °C with shaking (220 rpm) for 80 min in open–capped tubes and clarified by centrifuging twice at 12000 g 4 °C for 10 min. The resulting crude extract was aliquoted, flash frozen in liquid nitrogen and stored at –80 °C until use.

#### Isothermal Nucleic Acid Amplification

*<u>Homemade NASBA</u>* reactions were performed as following, with indicated changes in the main text and supplementary figures. In general, the buffer composition was 50 mM Tris-Cl at pH: 8.0, 50 mM KCl, 6 mM MgCl_2_, 2 mM spermidine, 10 mM DTT, 0.1 mg/mL BSA, 0.5% (v/v) Triton X-100 (prepared as 10x and diluted to 1x in final reaction). Final reaction mixture contained 1x reaction buffer, deoxyribonucleotide mix (1 mM each), ribonucleotide mix (2 mM each), forward and reverse primers ([final] = 1 µM each), 0.01 U of Yeast inorganic pyrophosphatase (YIPP, New England Biolabs), 0.4 U RiboLock RNase inhibitor (ThermoFisher), 0.08 U RNaseH (New England Biolabs), RevertAid Reverse Transcriptase 128 U (ThermoFisher), 32 U Hi^®^-T7 RNA polymerase (New England Biolabs), 500 nM random duplex primer pair, 1 µL of template RNA (varying amounts), 10% (v/v) DMSO, filled up to 10 µL of final volume with RNase/DNase-free water. If the reactions were performed with pre-incubation step at 65 °C for 3 min, the enzyme cocktail was assembled prior to the reaction and provided after the pre-incubation step.

*<u>Commercial NASBA</u>* kit was by AMS-Biotechnology (AMS.NLK.10). The NASBA reactions were assembled according to manufacturer’s instructions with following exceptions. The reaction mixture contained random primer duplex CF130/CF131 and the lyophilized mixture was rehydrated in final volume of 5 µL for each reaction, assembled with 1 µL of template RNA (added last). The reactions were performed at 41 °C for 90 min with 65 °C pre-incubation step for 3 min, and the enzymes were added after pre-incubation, as recommended by the manufacturer.

*<u>Commercial Reverse-Transcriptase RPA (RT-RPA)</u>* was by TwistAmp^®^ Basic Kit (TwistDx, UK). The reactions were assembled according to manufacturer’s recommendations with following exceptions. The total volume of each reaction was 10 µL. The RPA reactions contained 0.2 µL (40 U) RevertAid Reverse Transcriptase. For testing the clinical samples, the magnesium acetate (MgOAc) was <u>not</u> added last, instead, the template RNA was added last to start the reactions. The primers for all amplification methods are provided in Table S2.

*<u>Reverse-Transcriptase Rolling Circle Amplification (RT-RCA)</u>* was assembled in-house as following. Phi29 DNA polymerase (ThermoFisher) was provided at 10 U/µL, with the commercial buffer. The reaction mixture contained RevertAid Reverse Transcriptase (20 U) and T4 RNA Ligase 2 (10 U) or SplintR® RNA Ligase (10 U) with the splinting DNA oligonucleotide (ca. 22-25 bp overlaps) that is complementary to the 5’ and 3’ ends of the template RNA, which was 5’ UTR(+) sequence of SARS-CoV-2. The primers used for attempted amplification was given in Table S2. The total volume of each reaction was 10 µL.

#### *In vitro* transcription/translation

All *in vitro* transcription–translation reactions were performed in a final volume of 10.5 µL. In-solution reactions were recorded either in a Rotor–Gene Q qPCR machine (Qiagen) or in a multi-well plate reader (Varioskan, ThermoFisher). For plate reader experiments, 384-well black bottom plate (Corning) was used. BSA-blocked paper-disks were put in in clear bottom 96–well plate (Costar). For in-solution *E. coli* S12 reactions, the compositions were partially taken from the literature^37^ as following: amino acid solution (1.5 mM each) and energy solution (30 mM 3–phosphoglyceric acid, 1.5 mM ATP, 1.5 mM GTP, 0.9 mM CTP, 0.9 mM UTP, 0.2 mg/mL tRNA, 0.255 mM coenzyme A, 0.33 mM NAD^+^, 0.75 mM cAMP, 0.0675 mM folinic acid, 1 mM spermidine, 33 mM Na^+^–HEPES at pH 8.0.). The supplementary solution contained 2% (w/v) polyethyleneglycol 4000 (PEG 4000), 10 mM Mg^2+^–glutamate, 10 mM K^+^–glutamate, 20 mM sucrose, 12 mM maltose. For on-paper reactions, the mixture contained 50 ng/ µL purified and TEV-digested LacZω and β-galactosidase substrate Chlorophenol Red-β-D-galactopyranoside (CPRG, 0.12 µg/µL, Merck). Template plasmid DNA was obtained using Qiagen Plasmid Midi kit (12145) and used at a final concentration of 30 nM.

#### General Procedure for Colorimetric Assay

Prior to on-paper reaction, 1 µg of MBP-LacZω was digested by 1 µL TEV protease (New England Biolabs) in a 50 µL at 30 °C for 5 h. Upon completion of the digestion, the mixture was kept at 4°C (<12 h), until flash-freezing by liquid nitrogen. The paper disks were cut with an office paper hole punch, and blocked with 5% (w/v) BSA for 16 h with orbital shaking (50 rpm) at room temperature. Then, the paper disks were washed with RNase/DNase-free water 5 min for 3 times and let air-dry for 16 h. Prior to colorimetric assay, the paper-disks were put in in clear bottom 96–well plate (Costar) and the *E. coli* cell-free reactions were added on top (10 µL). The synthetic RNA was amplified by RT-RPA and added as 0.525 µL (1:20 dilution). Clinical RNA samples were either amplified by RT-RPA or magnetic-bead isolated and then amplified by RT-RPA and also provided as 0.525 µL (1:20 dilution). Then, the reactions were again let air-dry and monitored for color change at room temperature.

#### Synthetic and Clinical Sample Preparation

*<u>For synthetic sample preparation</u>*, the sequence of 5’ UTR(+) region of the wild-type SARS-CoV-2 (265 bp) was taken from the website of National Center for Biotechnology Information (NCBI, RefSeq: NC_045512.2). The sequence was synthesized as a dsDNA gene fragment from GenScript and cloned into an expression cassette (in plasmid pSB1A3) under the control of consensus T7 promoter, with additional 5’ flanking ATT and 3’ flanking GG sequences. *In vitro* transcription reaction was assembled in-house and performed by T7 RNA polymerase for 12 h at 37 °C. The final reaction mixture contained 100 µg/mL Bovine Serum Albumin Fraction V (BSA), 10 mM dithiothreitol (DTT), 2 mM ribonucleotide mix (each), 1 U Yeast Inorganic Pyrophosphatase, 0.4 U RiboLock RNase inhibitor (ThermoFisher), 500 U Hi^®^-T7 RNA polymerase (New England Biolabs), *in vitro* transcription buffer (1X: 100 mM K^+^-HEPES at pH 7.5, 5 mM MgCl_2_, 1 mM spermidine), and a column-purified template DNA PCR product (1 µg) in a total of 100 µL reaction volume filled-up by RNase/DNase-free water. Freshly synthesized RNAs were initially cleaned up by TRIzol and ethanol precipitated as following: acidification by 300 mM sodium acetate pH: 5.2, followed by 100% ice-cold ethanol, incubation at –20 °C for 2 h, centrifuged down at 15000 g for 30 min, and washed twice with 70% ethanol and cleaned by subsequent centrifugations. The air-dried pellets were resuspended in 50 µL water and further purified by NucleoSpin RNA Clean-up kit (Macherey-Nagel), aliquoted and dried by vacuum (CentriVap, Labconco) at ambient temperature for 16 h.

*<u>Clinical samples</u>*. The RNA from infected patients was derived from anonymized saliva samples collected with ethical committee approval of the Azienda Provinciale per i Servizi Sanitari of the Autonomous Province of Trento (P.A.T.). viral particles in the samples were inactivated by 6 M guanidinium chloride and RNA was purified with TRIzol reagent. (7 samples were used in this study)

*<u>For RNA isolation by magnetic beads</u>*, biotinylated DNA oligonucleotides (0.4–1 nmol, 25 bp) were complementary to 5’ UTR(+) region. Approximately 0.2–1 mg streptavidin-coated magnetic beads were used to capture the complementary RNA. For experiments used synthetic RNA spiking inside the saliva, the saliva samples were diluted 1:5 in phosphate-buffered saline (PBS) and cellular debris was cleared by centrifugation at 500 g for 5 min at room temperature. Subsequently, the RNA dilutions were spiked-in the sample. Prior to RNA isolation, magnetic beads were washed once and equilibrated for 10 min in high-salt binding buffer (50 mM Tris-Cl at pH: 7.5, 500 mM NaCl, 1 mM EDTA and 0.5% (v/v) Triton X-100). The separation was by a magnetic rack by Invitrogen. Then, the sample and magnetic beads were mixed and, hand-warmed for 1 min and let incubate at room temperature for 10 min without agitation. The magnetic beads were then captured out of the solution by magnets, and the beads were washed once with high-salt buffer without Triton X-100 and twice with ice-cold low-salt wash buffer (50 mM Tris-Cl at pH: 7.5, 150 mM NaCl, 1 mM EDTA). The captured RNA was then eluted by TE buffer (10 mM Tris-Cl, 1 mM EDTA at pH: 7.5) from the final magnetic bead slurry at 65–70 °C.

#### Quantitative Real-Time Reverse Transcriptase PCR (qRT-PCR)

Prior to qRT-PCR, complementary DNA (cDNA) was synthesized by reverse transcription with iScript™ cDNA synthesis kit (Bio-Rad). For clinical RNA samples, the template was 1 µL (in 20 µL total volume). For magnetic-bead isolated clinical RNA samples, the template was 14 µL (max. amount in 20 µL). qRT-PCR was performed by SsoAdvanced™ SYBR^®^ Supermix (BioRad), with primer pairs DT146/DT147 or DT152/DT153 (see Table S2). qPCR was run at Bio-Rad CFX96 Real-Time machine and acquisition with FAM channel. The PCR cycling program included initial denaturation for 30 sec at 95 °C, and cycling denaturation for 10 sec at 95 °C followed by annealing/extension for 30 sec at 60 °C that was for 40 cycles with plate reading at every cycle. At the end of the run, a melting curve was generated from 65 °C to 95 °C with 0.5°C/step increments. qRT-PCR primer pairs were designed by online software by MIT-Primer3 and Integrated DNA Technologies (IDT), optimized for T_m_ = 57–60 °C, to generate an amplicon size of 100– 130 bp. Standard curve was generated by serial dilutions of cDNA at 1:10 increments and primer efficiency was calculated as the slope of cycle threshold vs dilution factor. Efficiency between 90–110% was considered as acceptable.

#### Genetic Constructs and Protein Purification

*<u>Design of toehold switches:</u>* Toehold switches were designed by taking previously published reports as a guide^34,38^. In brief, the viral complementary sequences were joint with an 11-bp stem-region. The secondary structure predictions were obtained by NUPACK software (http://www.nupack.org/). Stem-region melting temperatures were calculated according to Primer3 software defaults (Santa Lucia 1998). The goal melting temperature was above 25 °C. The overall target Gibbs free energy of the secondary structure (ΔG°) was above –35 kcal/mol. In particular, complex secondary structures were avoided in the 25 bp toehold-flanking sequences.

All *<u>clonings</u>* were performed by in-house assembled Gibson Assembly mix^35^. All gene *<u>expression</u>* plasmids were with pSB1A3 backbone from iGEM Parts Registry. dsDNAs were obtained by PCR, gBlocks or in-house assembly of DNA primer-stitched templates. β-galactosidase ω-fragment (LacZω) was cloned into a modified pMAL-c4X backbone containing N-terminus Maltose Binding Protein (MBP) with TEV protease recognition site flanked by (GS)_2_ linker sequence. *E. coli* NEB5α strain transformed with MBP-TEV-LacZω expressing plasmid was grown in Terrific Broth at 37 °C. The fusion protein was overexpressed with Autoinduction Medium^36^ with overall growth of 24 h. The MBP-fusion protein was purified by Amylose Resin (New England Biolabs) according to manufacturer’s instructions and eluted by 10 mM maltose. After elution, first four fractions that contain the MBP-LacZω was pooled, flash frozen in liquid nitrogen and stored at –80 °C until use. The concentration of the proteins was determined by Pierce(tm) BCA Protein Assay Kit (ThermoFisher). The list of primers, genetic constructs and plasmids can be found at Table S2, including their sequences.

### Supplementary Figures

**Figure S1.**
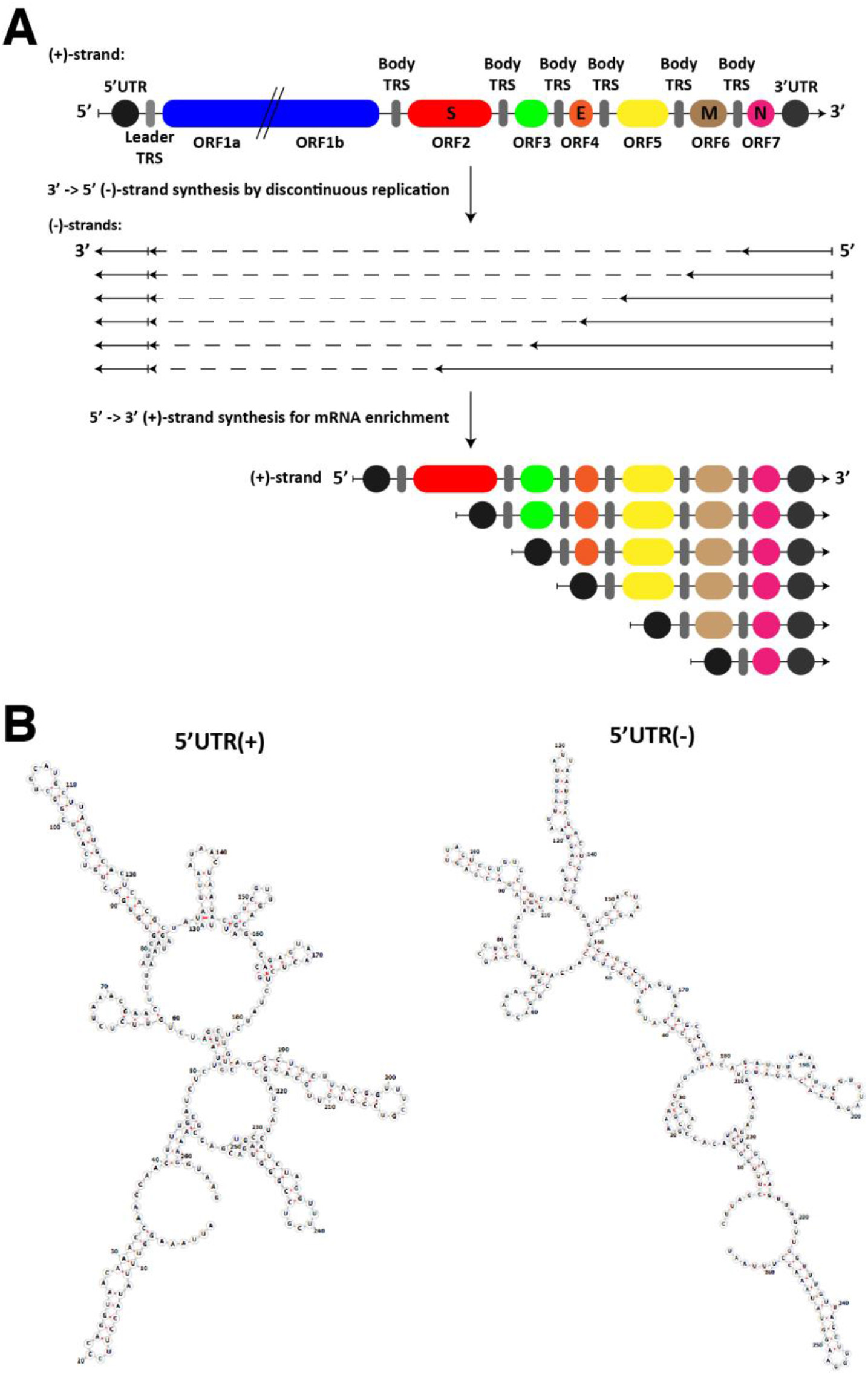
Overview of riboregulatory elements with respect to coronavirus replication cycle. **(A)** Discontinuous transcription of coronavirus genome results with 5’ and 3’ UTR enrichment in the synthesized RNAs. **(B)** Predicted secondary structures of 5’ UTR(+) and 5’UTR(−) strands of the SARS-CoV-2. Loop regions are predominantly chosen for complementary toehold sequences and stem regions are avoided in the design, as much as possible. The predictions were obtained by University of Vienna’s “RNAfold server” (http://rna.tbi.univie.ac.at/cgi-bin/RNAWebSuite/RNAfold.cgi).

**Figure S2.**
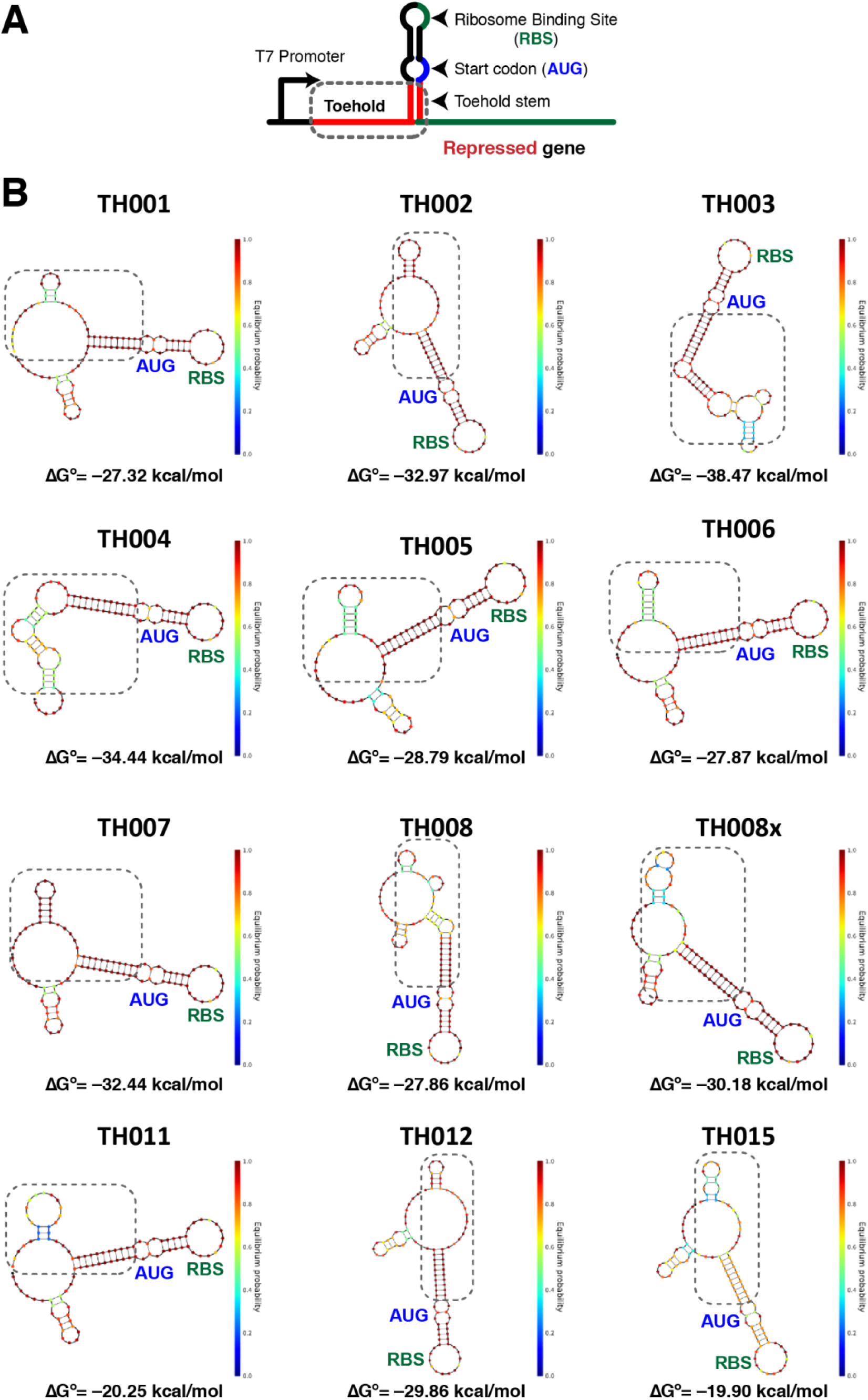
Design of different toehold-switch-mediated riboregulatory elements. The predicted secondary structures were obtained with NUPACK online software at 30 °C (http://www.nupack.org/).

**Figure S3.**
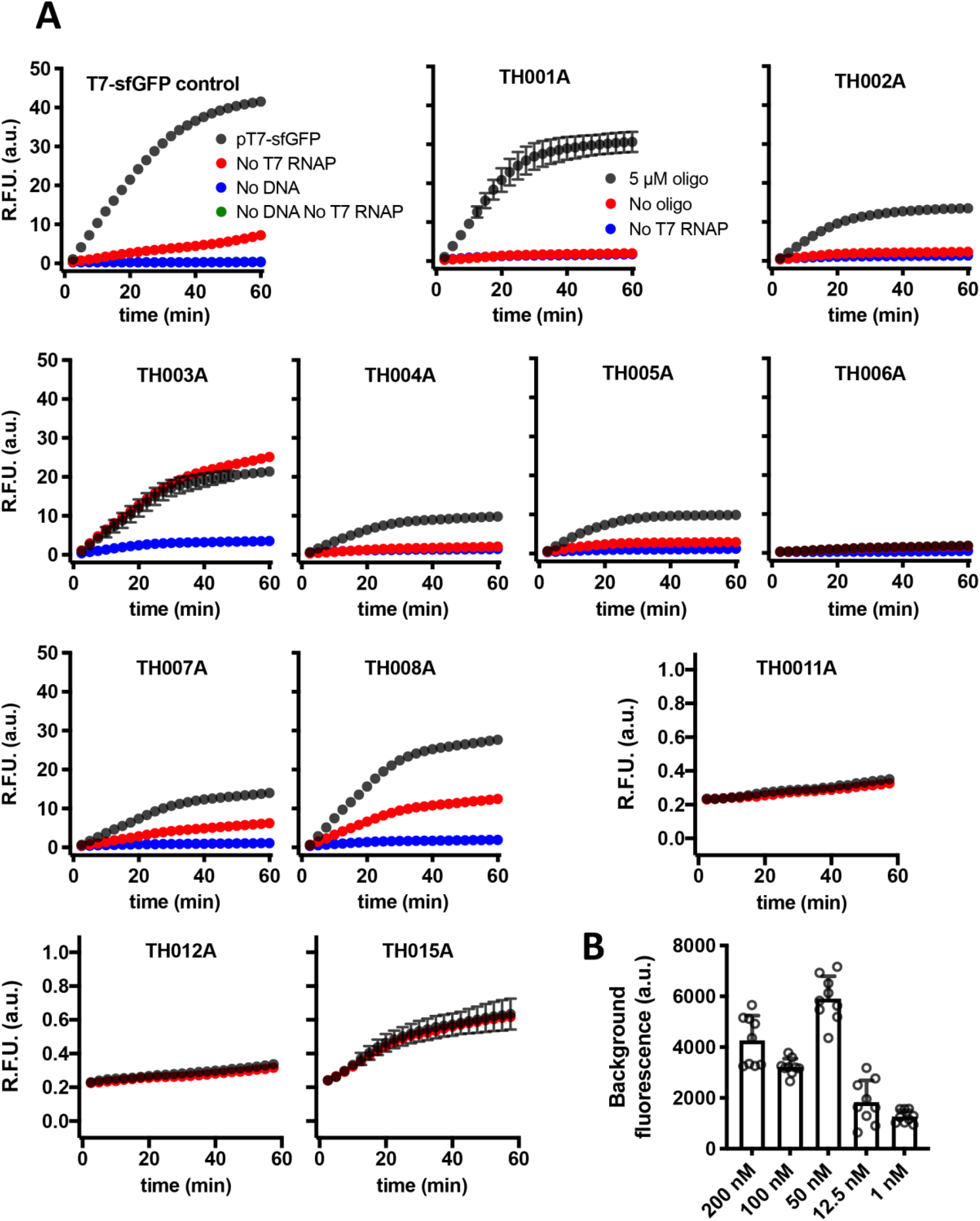
Functional screening of different toehold-switch-mediated riboregulatory elements. **(A)** Activities of different riboregulatory elements. **(B)** Minimum synthetic RNA concentrations that can activate gene expression in *E. coli* cell-free reaction. Note that 3’ UTR toehold-switches did not succeed (TH011A, TH012A, TH015A), likely due to highly stable secondary structures of target RNAs.

**Figure S4.**
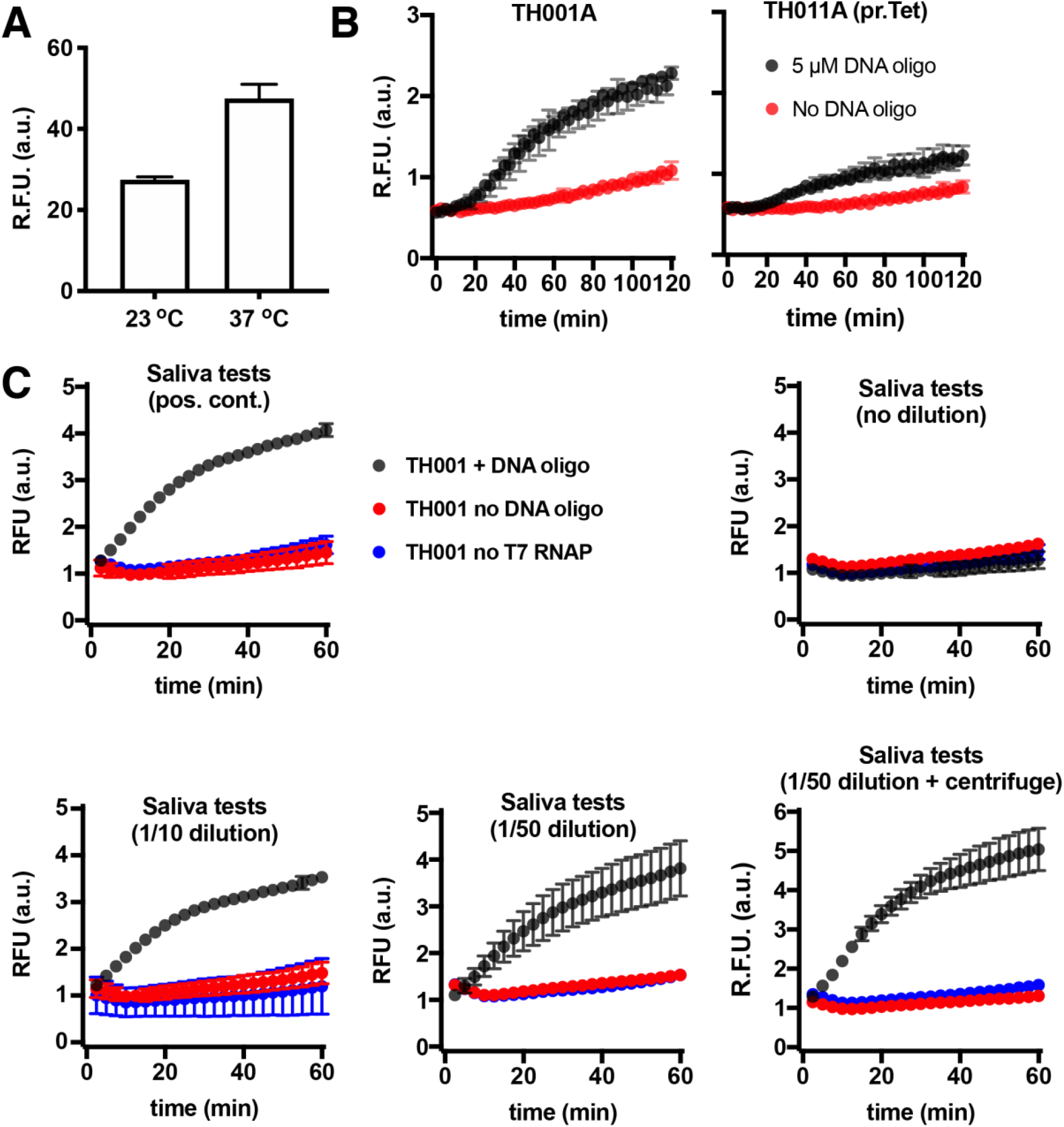
Optimization of *E. coli* cell-free protein expression system for RNA diagnostics. **(A)** Log-phase growth test. **(B)** Vacuum drying conditions of cell-free reactions. **(C)** Effect of saliva on cell-free reactions.

**Figure S5.**
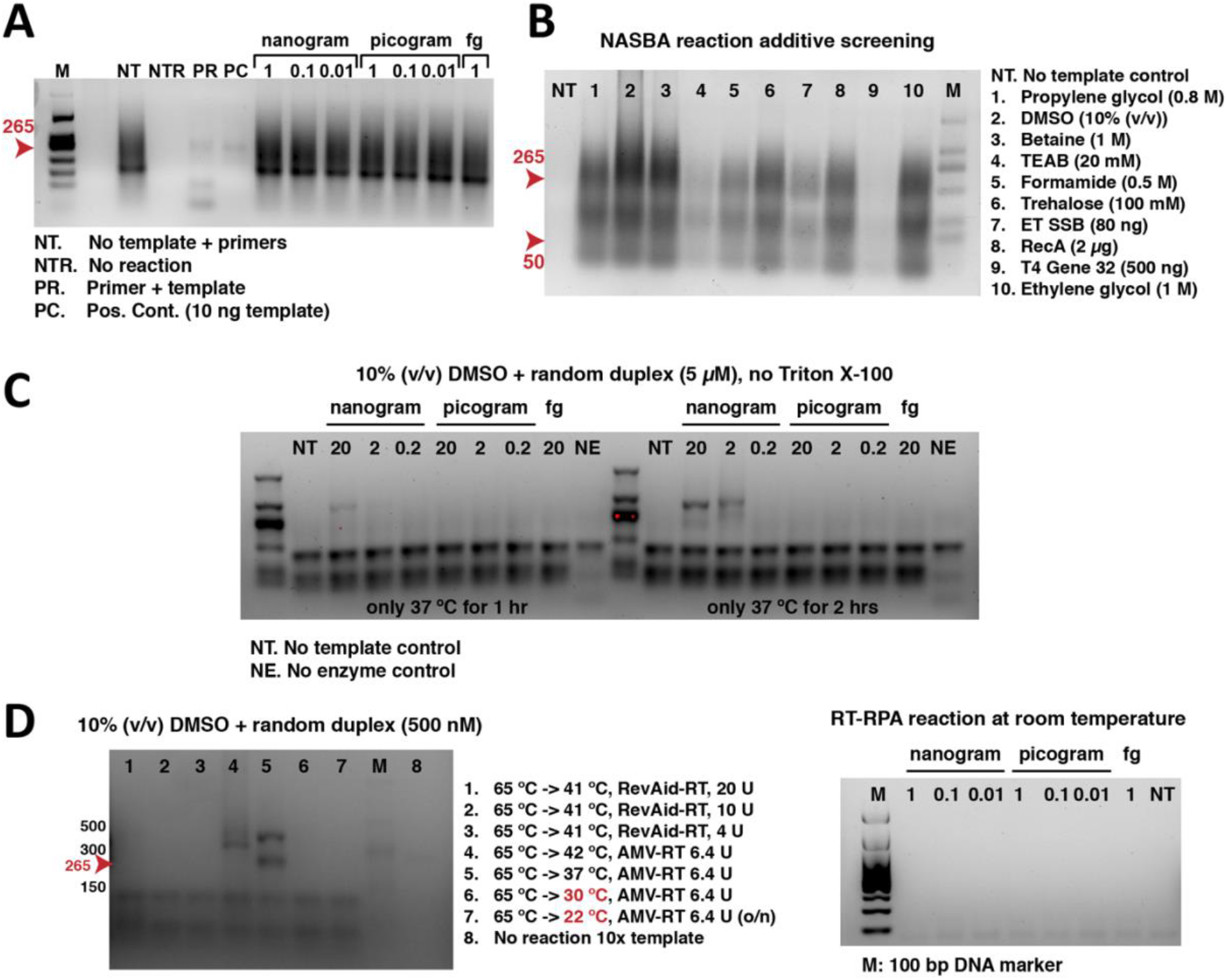
Optimization and screening of isothermal RNA amplification platforms. **(A)** Non-specific products were detected with NASBA reaction, in parallel with previous reports^30^ **(B)** Homemade NASBA reaction additive screening. **(C)** Influence of random RNA duplex on T7 RNA polymerase efficiency. Please note that more pronounced RNA bands were observed with the additions of the random duplex, in comparison to (A) and (B). **(D)** Influence of different reaction conditions and temperature cycling. **(D)** Room temperature amplifications by NASBA and RT-RPA. (Note that prior to loading onto gel, RT-RPA reactions –but not NASBA– were pre-incubated at 95 °C for 3 min and immediately placed on ice, in order to dissociate the SSB from the amplicons.) We also note that under reported reaction conditions, our initial attempts for RT-RCA did not yield amplification.

**Figure S6.**
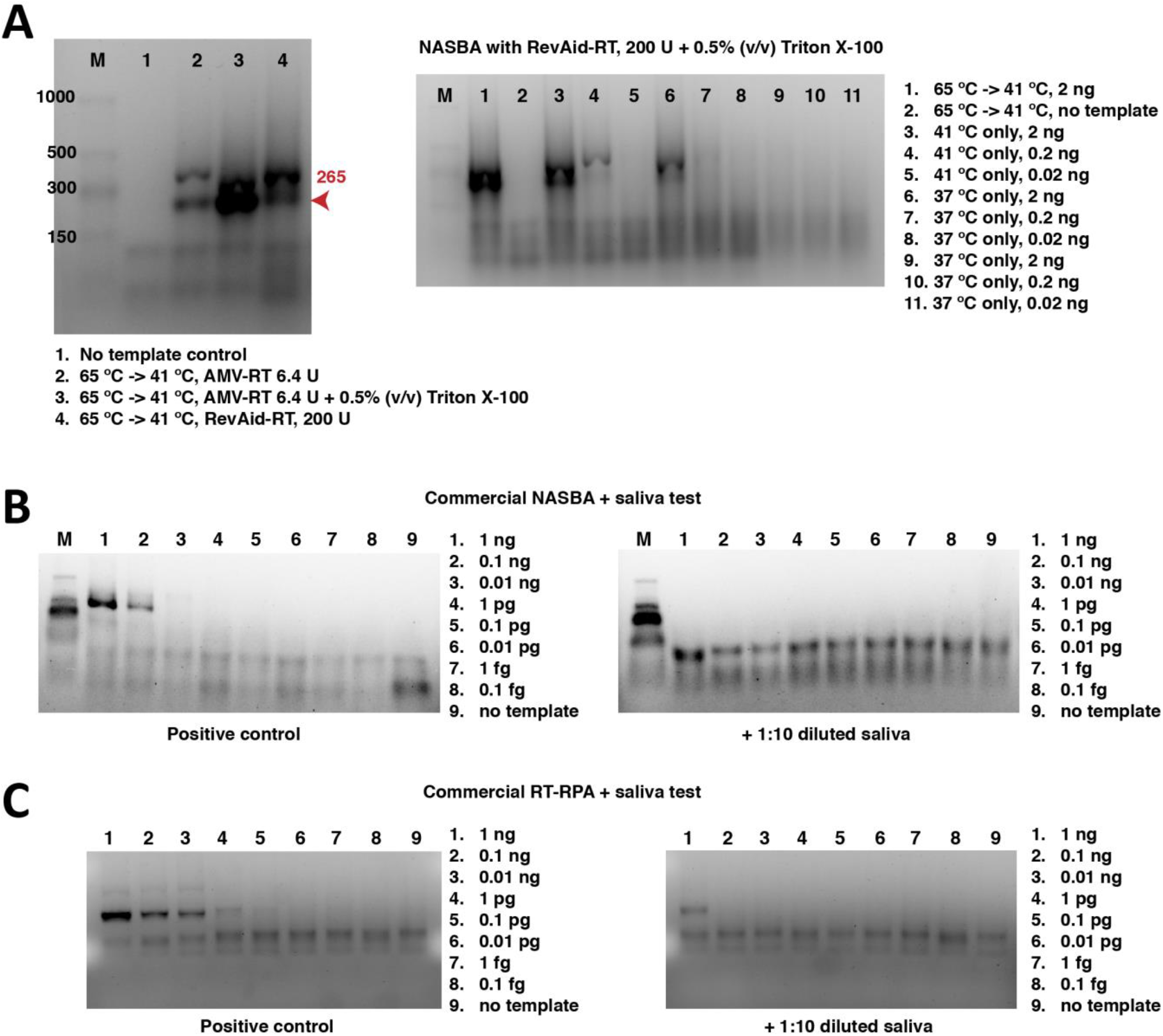
Testing compatibility of one-pot lysis buffer with isothermal RNA amplification. **(A)** 0.5% (v/v) Triton X-100 improved amplification efficiency in homemade NASBA mixture. **(B)** Homemade NASBA mixture was not conductive for one-pot cell lysis and RNA amplification. **(C)** RT-RPA with one-pot cell lysis buffer from saliva samples with RNA amplification at 37 °C.

**Figure S7.**
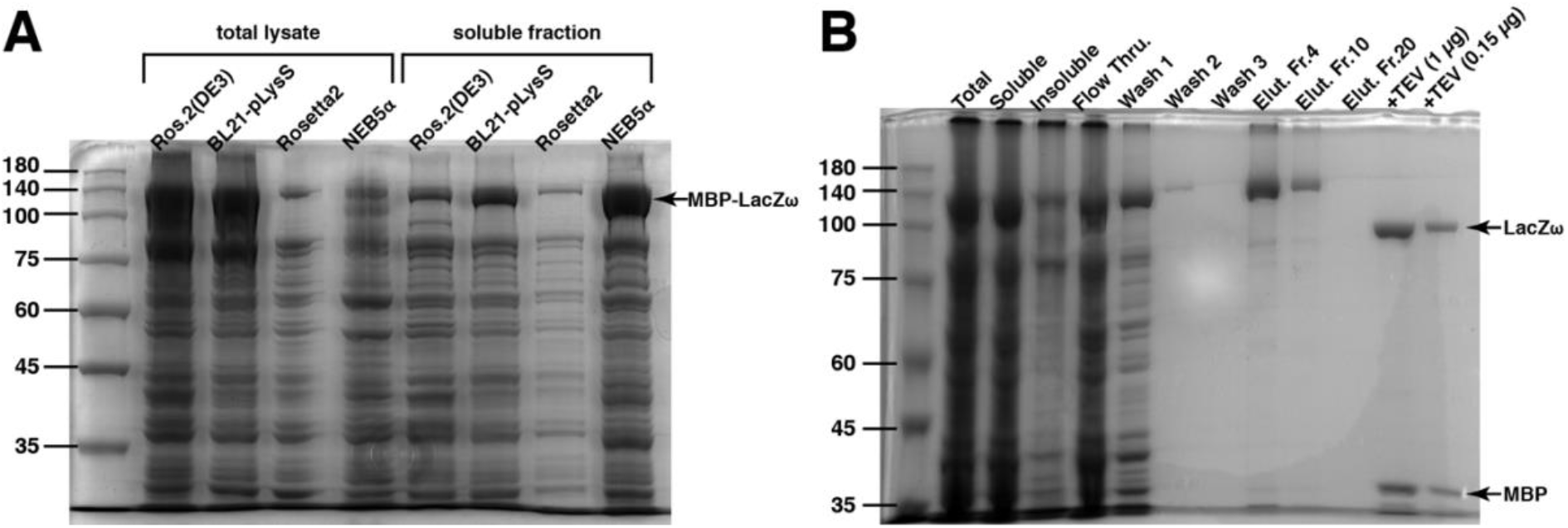
Expression and purification of β-galactosidase ω-fragment (LacZω). **(A)** Expression tests of different *E. coli* strains for Autoinduction medium at 24 h, 37 °C. **(B)** Purification and TEV protease digestion of MBP-LacZω. The molecular weight of LacZω is expected to be ca. 100 kDa and MBP (MalE gene product) is expected to be ca. 43 kDa.

**Figure S8.**
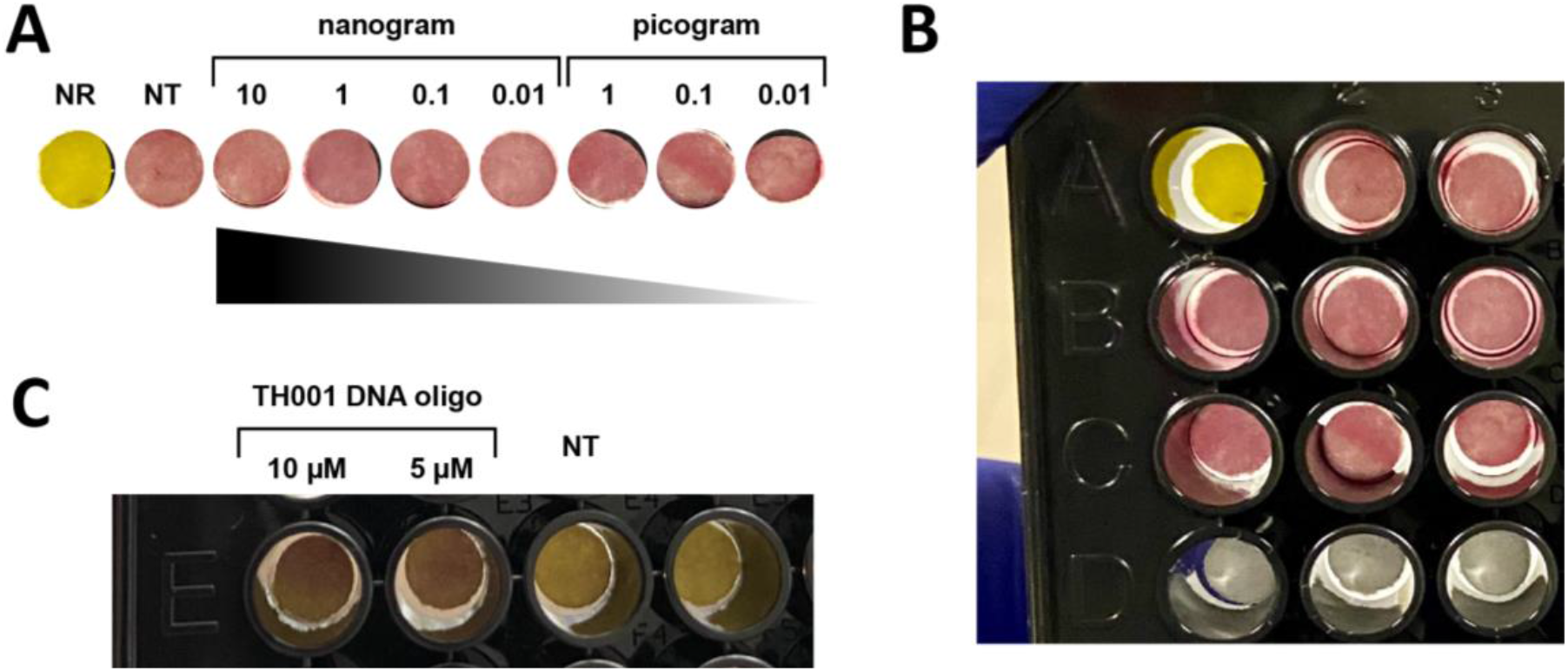
Characterization of *E. coli* cell-free protein expression systems for colorimetric output. **(A)** Rosetta 2(DE3) Singles strain endogenous LacZ activity with the colorimetric reporter. **(B)** Uncropped images. **(C)** Positive control of cell-free reaction colorimetric reporter, activated with complementary DNA oligonucleotide. NT stands for full reaction assembly but no template DNA oligonucleotide added to activate the cell-free gene expression.

**Figure S9.**
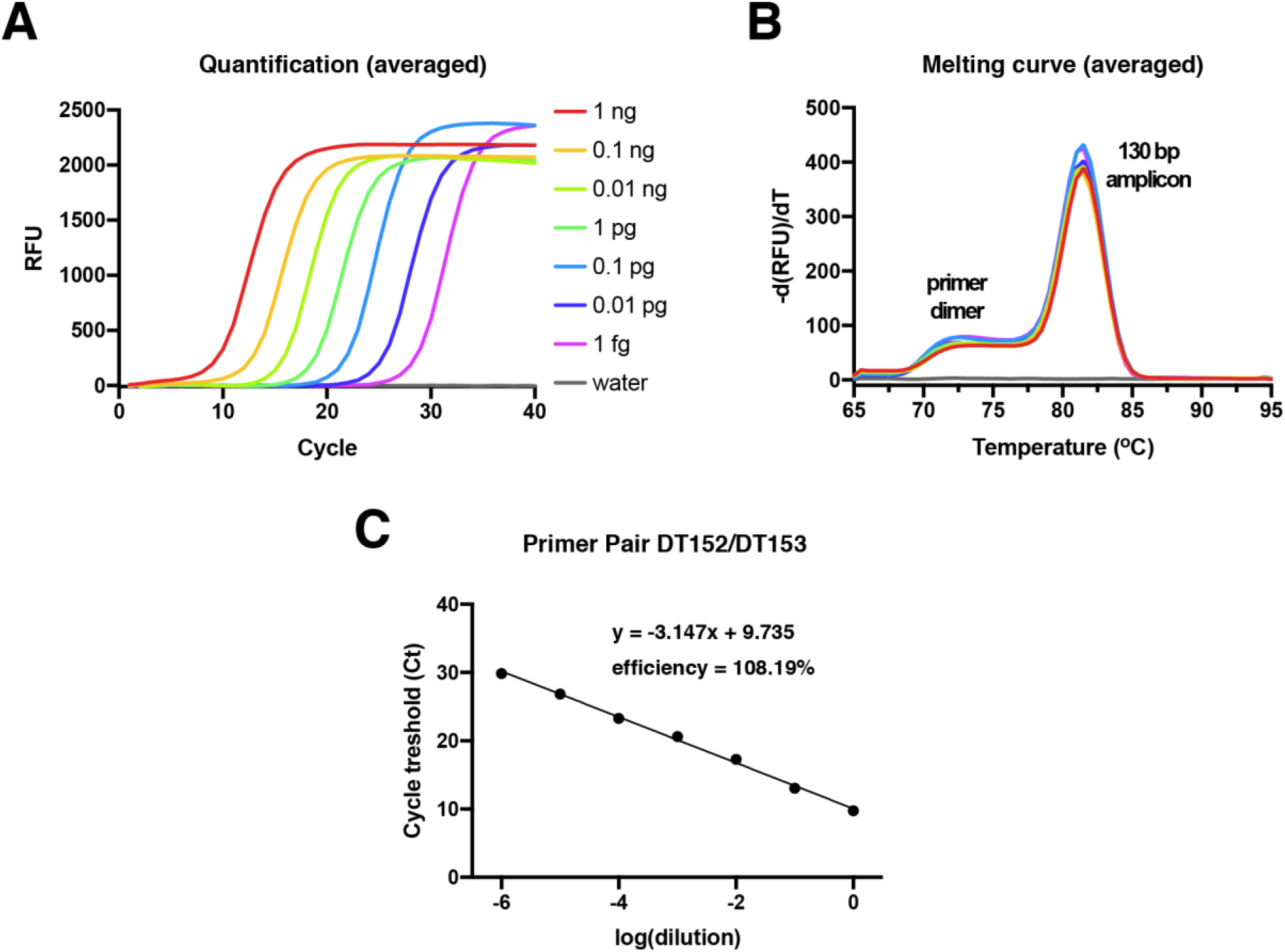
qRT-PCR verification of synthetic SARS-CoV-2 RNA [5’ UTR(+)] copy number. **(A)** Average quantification with serial dilution of cDNAs prepared from various concentrations of synthetic RNAs. This standard curve was used to determine the validity of RNA detection from clinical samples. **(B)** Melting curve analysis of the specific amplicon with the primer pair DT152/DT153. **(C)** The primer efficiency analysis of the pair. Note that primer pair efficiency of 90–110 % is accepted. By extrapolation of the standard curve, we determine minimum reliably detectable the copy number and the concentration of synthetic RNAs as 667 molecules and 100 attomolar (aM).

**Figure S10.**
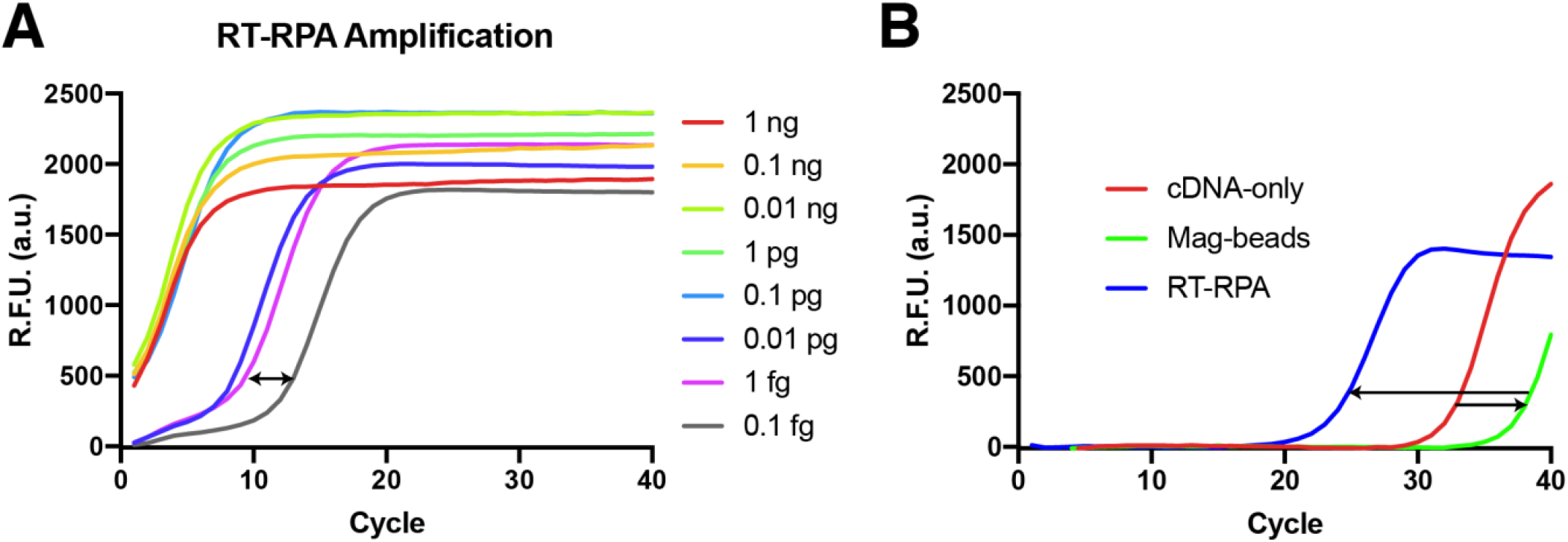
qRT-PCR verification of SARS-CoV-2 RNA [5’ UTR(+)] copy number from synthetic and clinical RNA. **(A)** Synthetic RNA detection after RT-RPA amplification. Range indicates potential colorimetric assay detection limit. **(B)** qRT-PCR verification of the clinical RNA. and saliva-spiked-in clinical RNA detection with magnetic bead isolation.

**Figure S11.**
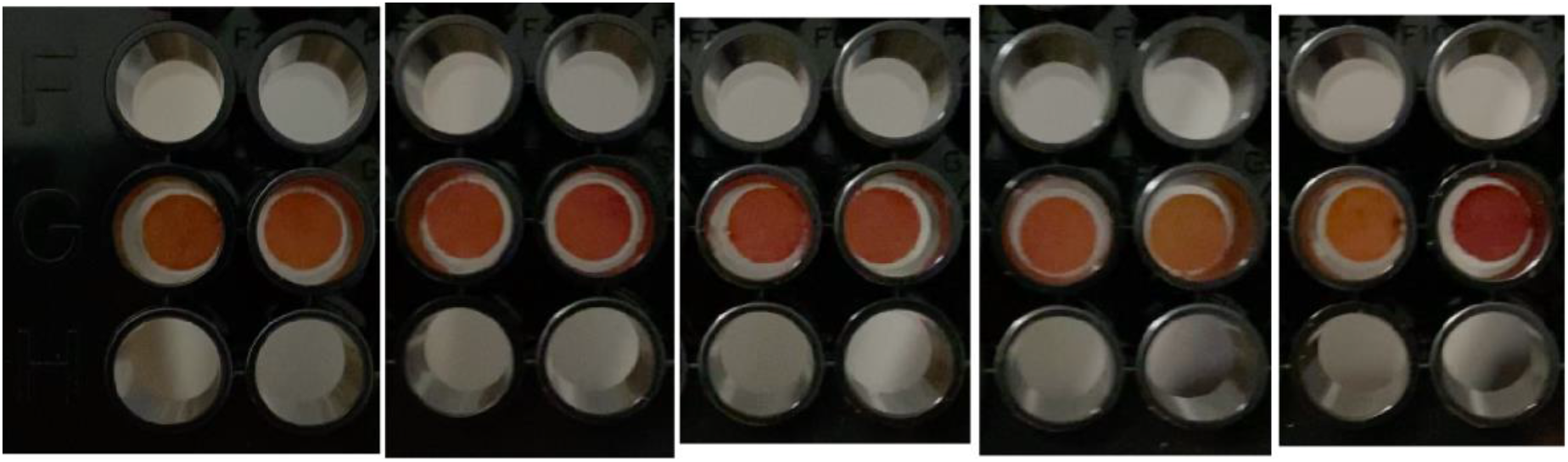
Plate pictures for Fig.1.

### Supplementary Tables

**Table S1.** Cost calculations and limitations. Comparative analysis of the *E. coli* cell-free protein expression system-based colorimetric RNA diagnostics with other approaches, with respect to commercial PURExpress, homemade OnePot PURE system^39,40^ and *E. coli* extracts^33,41^. All costs are provided in EUR (euro) currency (with 1.1xUSD=EUR estimate), per total reaction volume. For PURExpress, the reaction volumes are taken from ref.35, and for OnePot PURE, conventional commercial PURExpress volumes are taken as a reference, yet this is not supported by hard evidence in the literature. ***E. coli* extract:** We calculated the lysate preparation costs based on previous publications that were rigorously detailed^33,41^. In comparison to ref.33, we have not used any IPTG for induction and used ∼25% less extract (45% vs 33% of total volume), bringing an overall ∼20% reduction in costs, from ∼0.02 EUR/test (with extract as ∼50% of overall cost at ref.33) to 0.016 EUR/test. Here, we also note that in comparison to ref.40, the crude extract preparations require less purification steps, know-how and provides less room for unexpected failure in execution of protocols. Moreover, crude extracts also provide exceptionally convenient scale-up potential as opposed to resin-based purification systems such as OnePot PURE. **Energy, amino acid and other solutions:** The calculations were based on estimations of previous publications^33,39–41^. In comparison to ref.33, we have used ∼2-times less amino acid solution, decreasing our costs ∼10%, from ∼0.02 EUR/test to 0.018 EUR/test. The rest of the solution concentrations were comparable with similar breakeven costs. We also take into account that incurred plasmid preparation costs and general buffers/chemicals included in the reaction mixture. **RNA amplification**: Notably, we used an RT-RPA system, decreased down to 5 µL volume in a single proof-of-concept reaction, which did not require any additional incubators or thermocyclers. Given the amount used in the final test, which is 0.525 µL, the RT-RPA can also be performed in even smaller volumes such as 1 µL, reducing the associated costs further 5x-2.5x fold. 0.06-0.3 EUR/test for RPA elements. The cost of reverse transcriptase was 3 EUR/µL (200 U/µL) and we used 0.1 µL per 5 µL reaction (∼0.3 EUR/µL). In the end, for an averaged volume of 2.5 µL (5-times more than used final volume), the total cost estimate is ∼0.3 EUR/test (RPA: 0.15 EUR + RT: 0.15 EUR). The lowest end here would be ∼0.15 EUR/test, if 1 µL volume was used. **Colorimetric components:** To prepare LacZω protein, MBP-based purification (Amylose resin) components are roughly estimated to have 100 EUR per single purification, which typically yields 15 mg of total protein (and can be reused up to 5 times). 15 mg is enough for 300 tests (for 50 µg/reaction), with final cost of 0.33 EUR/test; (with commercial TEV protease used at 0.035 EUR/test). Overall cost can be reduced down to ca. 0.033 EUR/test (10-fold), if one would like to adapt Ni^2+^-NTA-based purification (IMAC systems) instead of Amylose resin. Overall mix of colorimetric components were added as 0.525 µL in 10 µL test (1:20 dilution). Further, the MBP-LacZω can be digested in bulk, and at higher concentrations (we tested 4x more) and flash-frozen in liquid N_2_, and thawed for bulk testing at once. Taken altogether, the cost of colorimetric components is estimated as 0.36 EUR/test, with a lowest end estimated to be ∼0.068 EUR/test). **Labor:** The labor is considered from active work time as 0.227 EUR/µL reaction^40^. Generally, we reduced the lysate preparation times and costs by omitting the dialysis step (e.g., 1 h less active time) and adapted more streamlined workplan with ∼5 h of active time, provided no laborious purification steps are needed as opposed to OnePot PURE (additional ∼2 h active time deducted from 8 h estimate)^39,40^. Including all culture growth phases, the lysate preparation takes 1.5 labor days. We assumed assembly (actual test) duration is same for all, ∼1 h, totaling of ∼6 h active time. If one would like to make a total time estimate, we can conclude that our assay can be finished in less than 36 h from the first inoculation. **Magnetic beads (optional):** Biotinylated-oligonucleotide requires 100-500 pmol per capture, at a cost of ca. 1.5 euro/100 pmol. Streptavidin-coated magnetic beads are used at 100-500 µg per reaction (2-10 euro/sample). Nevertheless, this step is optional and subject to further optimization for reduction in mass of beads used; thus, this cost is not essentially relevant to the final cost estimation of our assay.

| Component | PURExpress<br>(commercial, per 2 μL) | OnePot PURE system<br>(homemade, per 2 μL) | *E. coli* S12 extract<br>(crude, per 10 μL) |
| --- | --- | --- | --- |
| Cell lysate or<br>enzyme solution | Sol. B: ~2 EUR | ~0.013 EUR | ~0.016 EUR |
| Energy, amino acid and<br>other solutions (total mix) | Sol. A: ~1 EUR | ~0.004 EUR | ~0.018 EUR |
| Colorimetric components<br>(LacZω + TEV + CRPG) | ~0.36 EUR (low: ~0.068<br>EUR) | ~0.36 EUR (not verified) | ~0.36 EUR (low: ~0.068<br>EUR) |
| RPA components +RT<br>(2.5 μL total volume) | ~0.3 EUR (low: ~0.12<br>EUR) | ~0.3 EUR (not verified) | ~0.3 EUR (low: ~0.12<br>EUR) |
| Labor | Active time ~2h: 0.0027<br>EUR (for test only) | Active time ~9 h: 0.012<br>EUR (for preparation + test) | Active time ~6 h: 0.008<br>EUR (for preparation + test) |
| Total | ~3.67 EUR <u>for 2 μL</u> | ~0.69 EUR <u>for 2 μL</u> | ~0.72 EUR <u>for 10 μL</u><br>(as low as ~0.23 EUR/10<br>μL<br>and ~0.023 EUR/μL) |

**Table S2.**
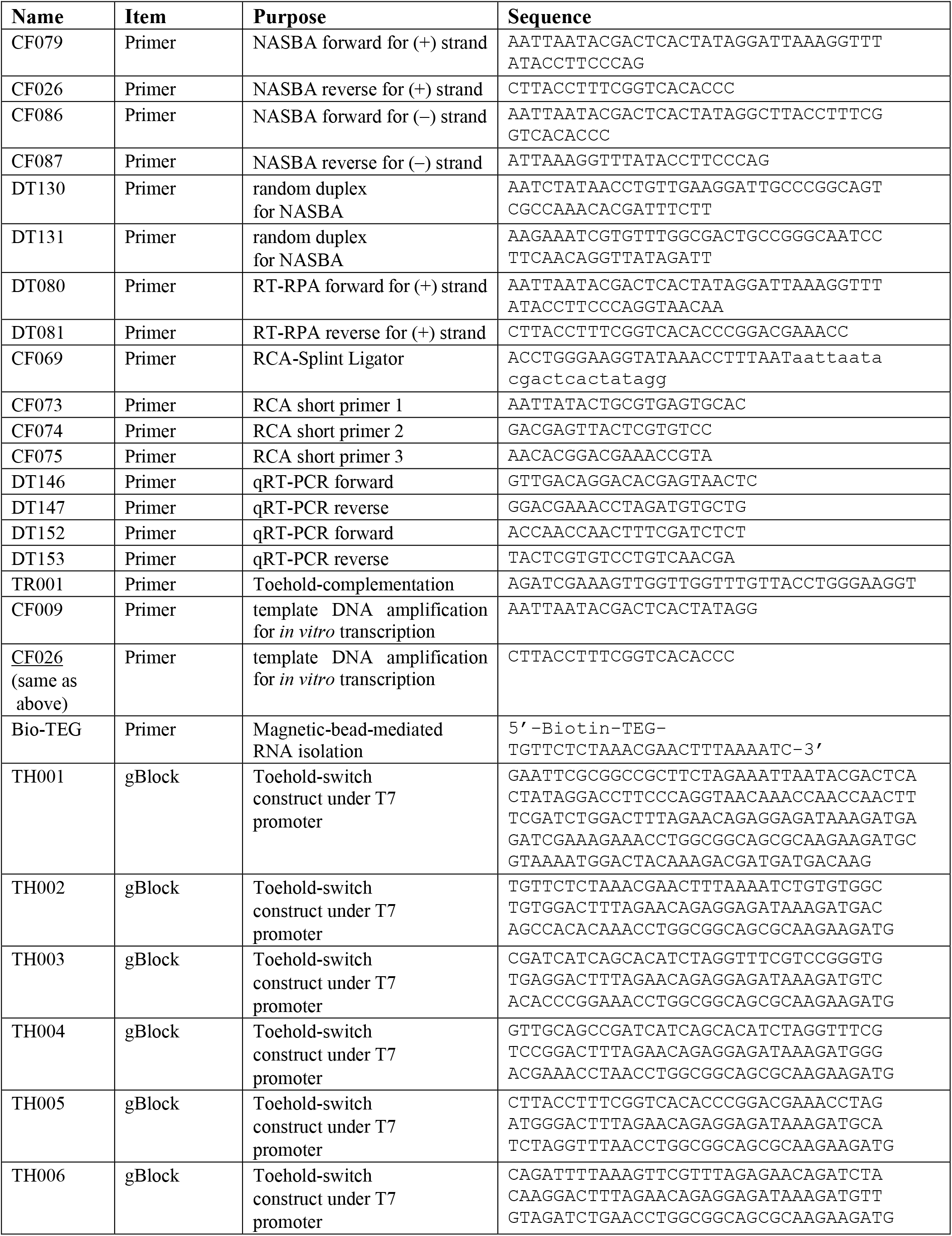
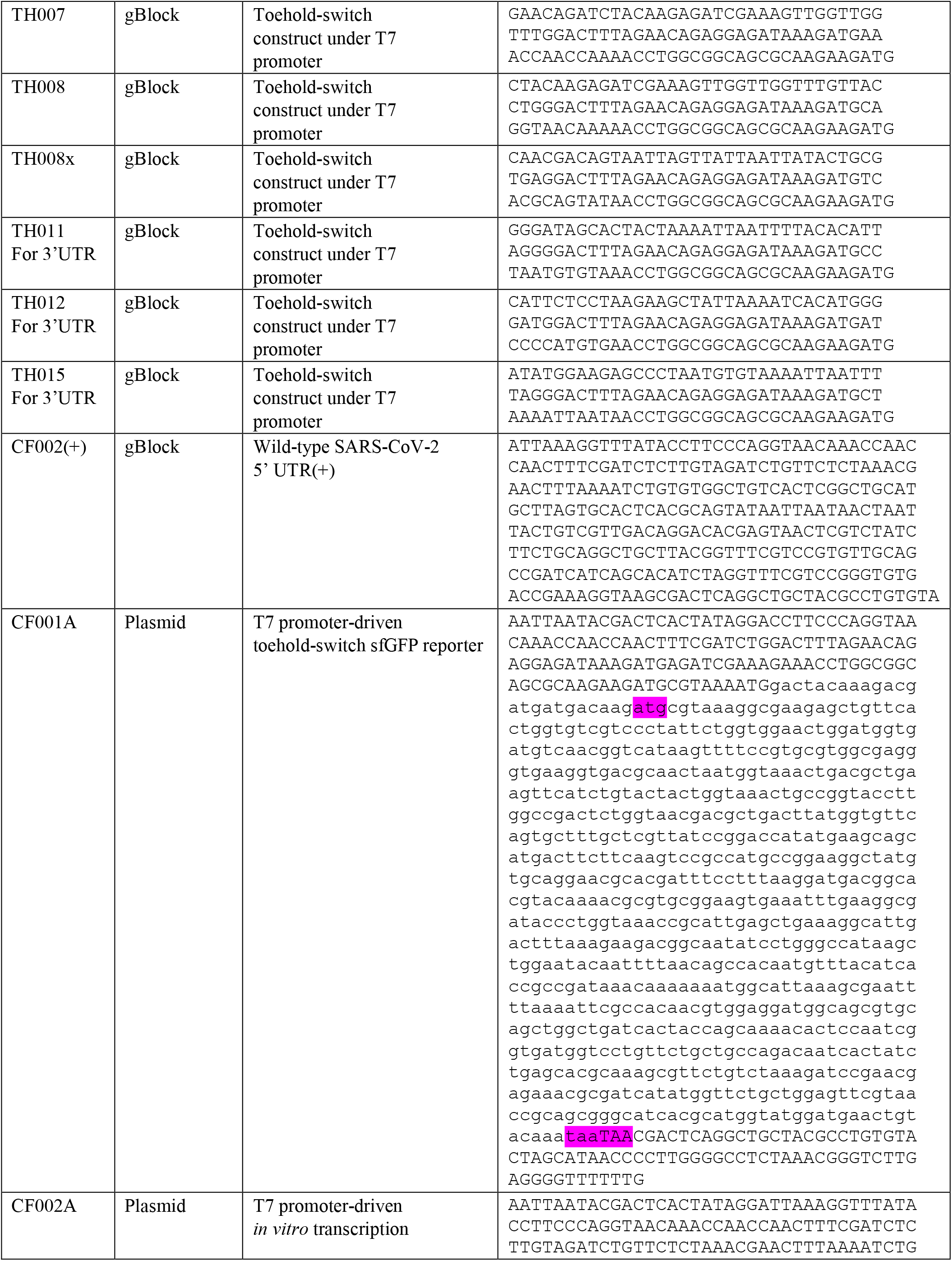
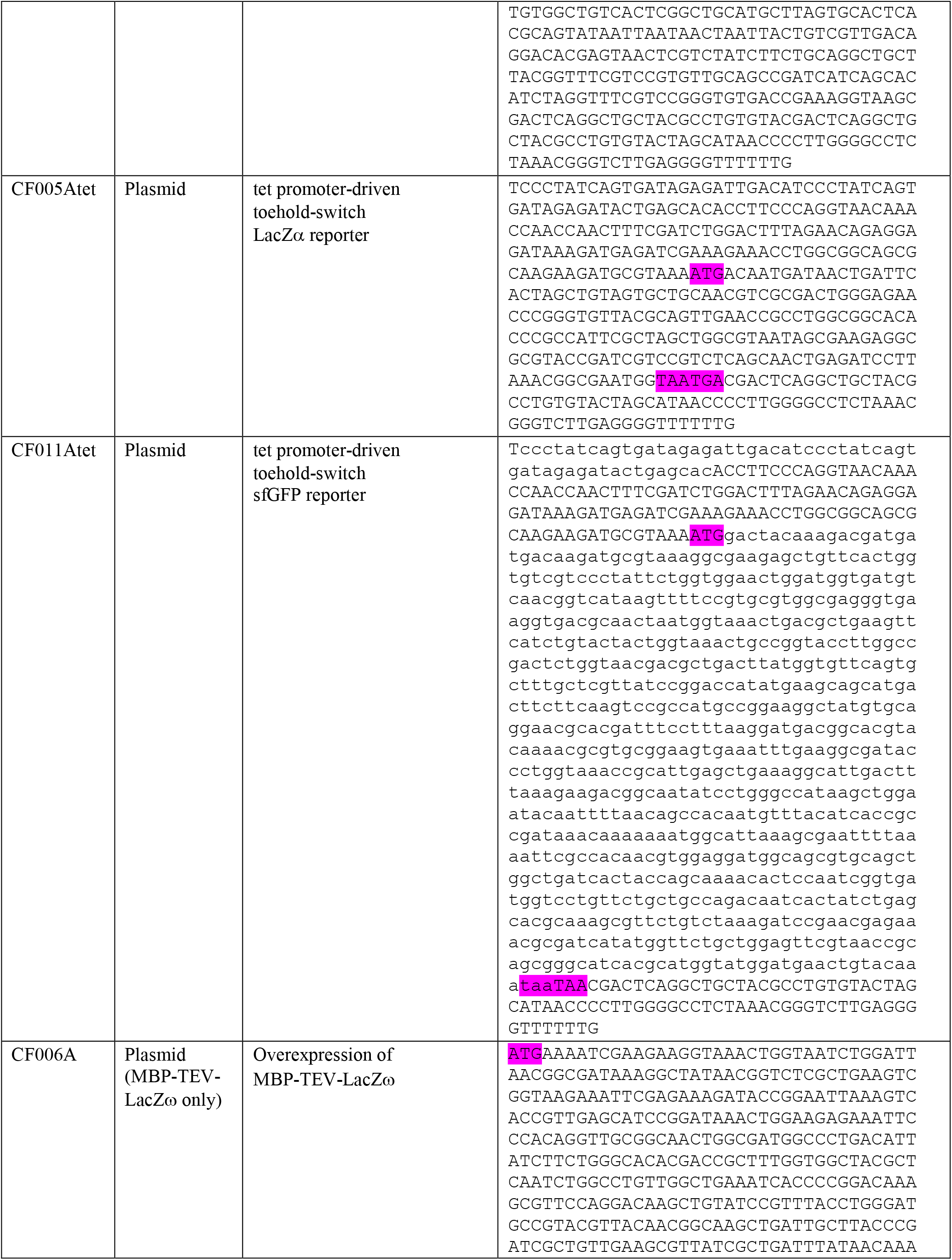
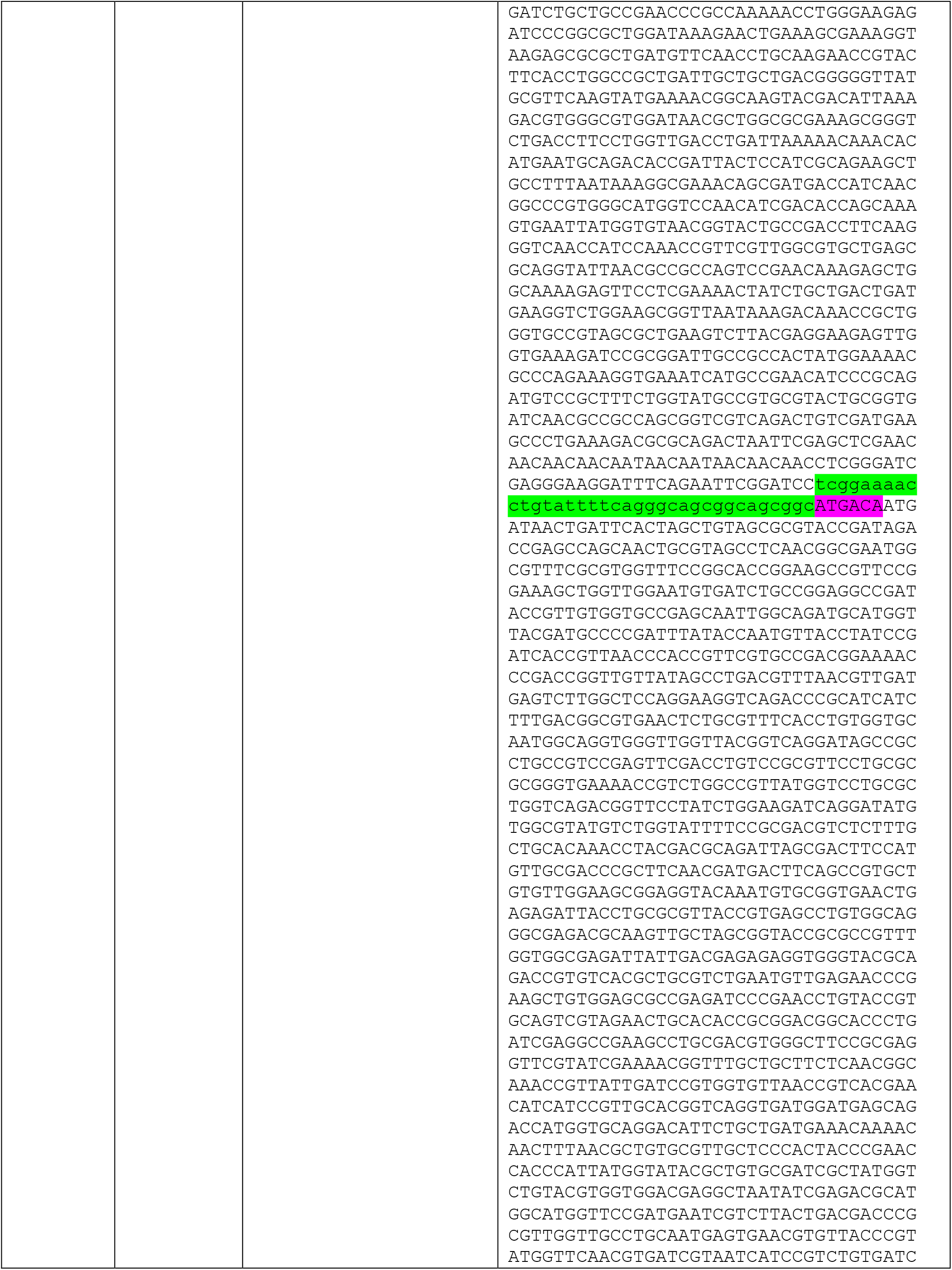
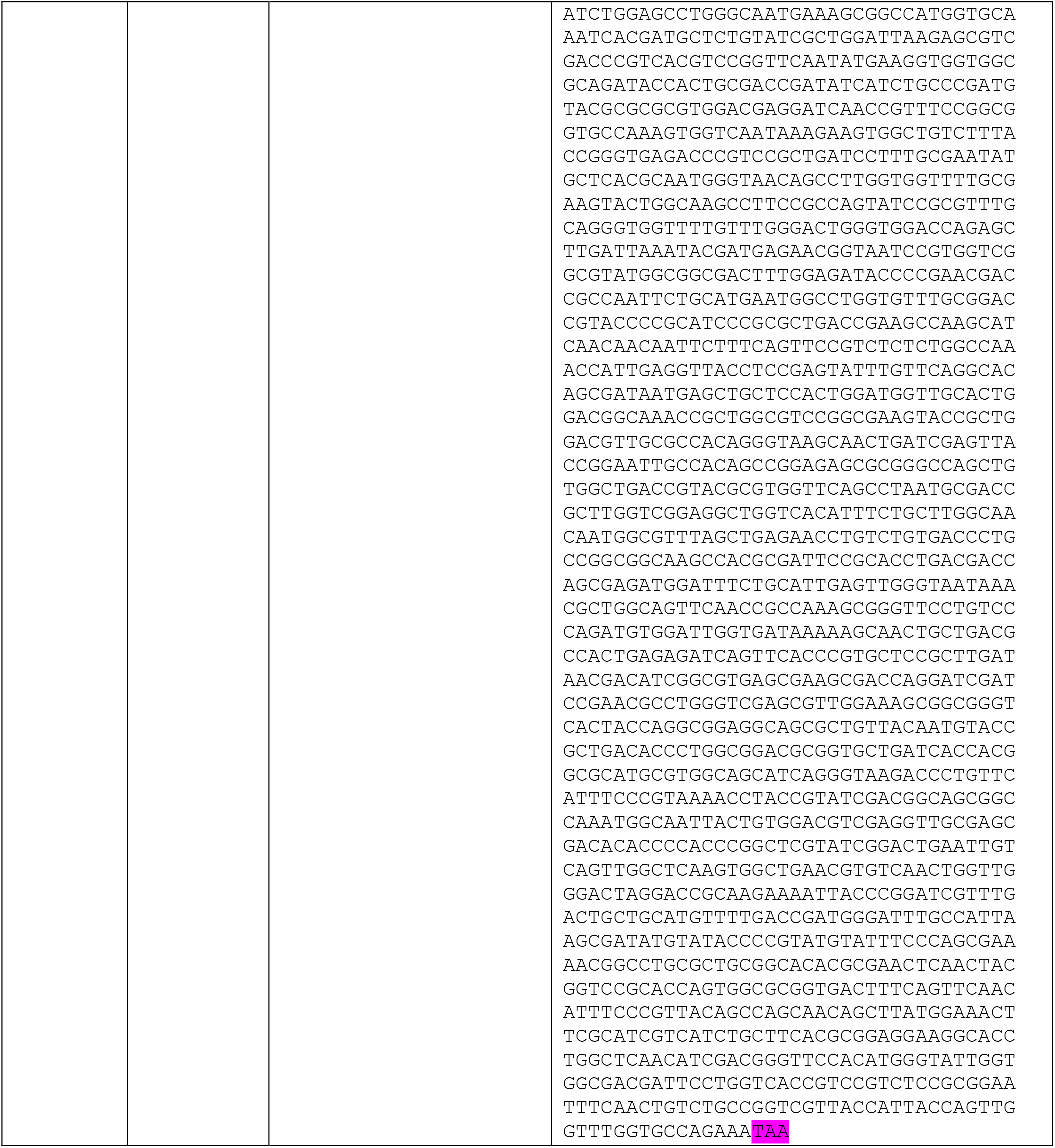
List of primers, genetic constructs and plasmids used in this study. Plasmid sequences are given without backbone. Start codons, stop codons and TEV protease recognition site (along with flexible linker) are highlighted in purple and green, respectively.

## REFERENCES

(1) Won, J.; Lee, S.; Park, M.; Kim, T. Y.; Park, M. G.; Choi, B. Y.; Kim, D.; Chang, H.; Kim, V. N.; Lee, C. J. Development of a Laboratory-Safe and Low-Cost Detection Protocol for SARS-CoV-2 of the Coronavirus Disease 2019 (COVID-19). Exp. Neurobiol. 2020, 29 (2), 1–13. https://doi.org/10.5607/en20009.

(2) Ding, X.; Yin, K.; Li, Z.; Liu, C. All-in-One Dual CRISPR-Cas12a (AIOD-CRISPR) Assay: A Case for Rapid, Ultrasensitive and Visual Detection of Novel Coronavirus SARS-CoV-2 and HIV Virus. bioRxiv 2020, 2020.03.19.998724. https://doi.org/10.1101/2020.03.19.998724.

(3) Carter, L. J.; Garner, L. V.; Smoot, J. W.; Li, Y.; Zhou, Q.; Saveson, C. J.; Sasso, J. M.; Gregg, A. C.; Soares, D. J.; Beskid, T. R.; Jervey, S. R.; Liu, C. Assay Techniques and Test Development for COVID-19 Diagnosis. ACS Cent. Sci. 2020, 6 (5), 591–605. https://doi.org/10.1021/acscentsci.0c00501.

(4) Pardee, K.; Green, A. A.; Takahashi, M. K.; Braff, D.; Lambert, G.; Lee, J. W.; Ferrante, T.; Ma, D.; Donghia, N.; Fan, M.; Daringer, N. M.; Bosch, I.; Dudley, D. M.; O’Connor, D. H.; Gehrke, L.; Collins, J. J. Rapid, Low-Cost Detection of Zika Virus Using Programmable Biomolecular Components. Cell 2016, 165 (5), 1255–1266. https://doi.org/10.1016/j.cell.2016.04.059.

(5) Slomovic, S.; Pardee, K.; Collins, J. J. Synthetic Biology Devices for in Vitro and in Vivo Diagnostics. Proc. Natl. Acad. Sci. U. S. A. 2015, 112 (47), 14429–14435. https://doi.org/10.1073/pnas.1508521112.

(6) Green, A. A.; Silver, P. A.; Collins, J. J.; Yin, P. Toehold Switches: De-Novo-Designed Regulators of Gene Expression. Cell 2014, 159 (4), 925–939. https://doi.org/10.1016/j.cell.2014.10.002.

(7) Pardee, K.; Green, A. A.; Ferrante, T.; Cameron, D. E.; Daleykeyser, A.; Yin, P.; Collins, J. J. Paper-Based Synthetic Gene Networks. Cell 2014, 159 (4), 940–954. https://doi.org/10.1016/j.cell.2014.10.004.

(8) Pardee, K.; Slomovic, S.; Nguyen, P. Q.; Lee, J. W.; Donghia, N.; Burrill, D.; Ferrante, T.; McSorley, F. R.; Furuta, Y.; Vernet, A.; Lewandowski, M.; Boddy, C. N.; Joshi, N. S.; Collins, J. J. Portable, On-Demand Biomolecular Manufacturing. Cell 2016, 167 (1), 248–259.e12. https://doi.org/10.1016/j.cell.2016.09.013.

(9) Silverman, A. D.; Karim, A. S.; Jewett, M. C. Cell-Free Gene Expression: An Expanded Repertoire of Applications. Nat. Rev. Genet. 2020, 21 (3), 151–170. https://doi.org/10.1038/s41576-019-0186-3.

(10) Rasor, B. J.; Vögeli, B.; Landwehr, G. M.; Bogart, J. W.; Karim, A. S.; Jewett, M. C. Toward Sustainable, Cell-Free Biomanufacturing. Curr. Opin. Biotechnol. 2021, 69, 136–144. https://doi.org/10.1016/j.copbio.2020.12.012.

(11) Fozouni, P.; Son, S.; Díaz de León Derby, M.; Knott, G. J.; Gray, C. N.; D’Ambrosio, M. V.; Zhao, C.; Switz, N. A.; Kumar, G. R.; Stephens, S. I.; Boehm, D.; Tsou, C. L.; Shu, J.; Bhuiya, A.; Armstrong, M.; Harris, A. R.; Chen, P. Y.; Osterloh, J. M.; Meyer-Franke, A.; Joehnk, B.; Walcott, K.; Sil, A.; Langelier, C.; Pollard, K. S.; Crawford, E. D.; Puschnik, A. S.; Phelps, M.; Kistler, A.; DeRisi, J. L.; Doudna, J. A.; Fletcher, D. A.; Ott, M. Amplification-Free Detection of SARS-CoV-2 with CRISPR-Cas13a and Mobile Phone Microscopy. Cell 2021, 184 (2), 323–333.e9. https://doi.org/10.1016/j.cell.2020.12.001.

(12) Joung, J.; Ladha, A.; Saito, M.; Kim, N.-G.; Woolley, A. E.; Segel, M.; Barretto, R. P. J.; Ranu, A.; Macrae, R. K.; Faure, G.; Ioannidi, E. I.; Krajeski, R. N.; Bruneau, R.; Huang, M.-L. W.; Yu, X. G.; Li, J. Z.; Walker, B. D.; Hung, D. T.; Greninger, A. L.; Jerome, K. R.; Gootenberg, J. S.; Abudayyeh, O. O.; Zhang, F. Detection of SARS-CoV-2 with SHERLOCK One-Pot Testing. N. Engl. J. Med. 2020, 383 (15), 1492–1494. https://doi.org/10.1056/NEJMc2026172.

(13) Mahas, A.; Wang, Q.; Marsic, T.; Mahfouz, M. M. A Novel Miniature CRISPR-Cas13 System for SARS-CoV-2 Diagnostics. ACS Synth. Biol. 2021. https://doi.org/10.1021/acssynbio.1c00181.

(14) Broughton, J. P.; Deng, X.; Yu, G.; Fasching, C. L.; Servellita, V.; Singh, J.; Miao, X.; Streithorst, J. A.; Granados, A.; Sotomayor-Gonzalez, A.; Zorn, K.; Gopez, A.; Hsu, E.; Gu, W.; Miller, S.; Pan, C. Y.; Guevara, H.; Wadford, D. A.; Chen, J. S.; Chiu, C. Y. CRISPR–Cas12-Based Detection of SARS-CoV-2. Nat. Biotechnol. 2020, 38 (7), 870–874. https://doi.org/10.1038/s41587-020-0513-4.

(15) Yan, L.; Zhou, J.; Zheng, Y.; Gamson, A. S.; Roembke, B. T.; Nakayama, S.; Sintim, H. O. Isothermal Amplified Detection of DNA and RNA. Mol. Biosyst. 2014, 10 (5), 970–1003. https://doi.org/10.1039/c3mb70304e.

(16) Yang, Q.; Meyerson, N. R.; Clark, S. K.; Paige, C. L.; Fattor, W. T.; Gilchrist, A. R.; Barbachano-Guerrero, A.; Healy, B. G.; Worden-Sapper, E. R.; Wu, S. S.; Muhlrad, D.; Decker, C. J.; Saldi, T. K.; Lasda, E.; Gonzales, P. K.; Fink, M. R.; Tat, K. L.; Hager, C. R.; Davis, J. C.; Ozeroff, C. D.; Brisson, G. R.; McQueen, M. B.; Leinwand, L.; Parker, R.; Sawyer, S. L. Saliva Twostep for Rapid Detection of Asymptomatic Sars-Cov-2 Carriers. Elife 2021, 10, 1–28. https://doi.org/10.7554/eLife.65113.

(17) Nguyen, P. Q.; Soenksen, L. R.; Donghia, N. M.; Angenent-Mari, N. M.; de Puig, H.; Huang, A.; Lee, R.; Slomovic, S.; Galbersanini, T.; Lansberry, G.; Sallum, H. M.; Zhao, E. M.; Niemi, J. B.; Collins, J. J. Wearable Materials with Embedded Synthetic Biology Sensors for Biomolecule Detection. Nat. Biotechnol. 2021. https://doi.org/10.1038/s41587-021-00950-3.

(18) Sola, I.; Almazán, F.; Zúñiga, S.; Enjuanes, L. Continuous and Discontinuous RNA Synthesis in Coronaviruses. Annu. Rev. Virol. 2015, 2 (1), 265–288. https://doi.org/10.1146/annurev-virology-100114-055218.

(19) Taiaroa, G.; Rawlinson, D.; Featherstone, L.; Pitt, M.; Caly, L.; Druce, J.; Purcell, D.; Harty, L.; Tran, T.; Roberts, J.; Catton, M.; Williamson, D.; Coin, L.; Duchene, S. Direct RNA Sequencing and Early Evolution of SARS-CoV-2. bioRxiv 2020, 2, 2020.03.05.976167. https://doi.org/10.1101/2020.03.05.976167.

(20) Kim, D.; Lee, J. Y.; Yang, J. S.; Kim, J. W.; Kim, V. N.; Chang, H. The Architecture of SARS-CoV-2 Transcriptome. Cell 2020, 181 (4), 914–921.e10. https://doi.org/10.1016/j.cell.2020.04.011.

(21) Hunt, J. P.; Zhao, E. L.; Free, T. J.; Soltani, M.; Warr, C. A.; Benedict, A. B.; Takahashi, M. K.; Griffitts, J. S.; Pitt, W. G.; Bundy, B. C. Towards Detection of SARS-CoV-2 RNA in Human Saliva: A Paper-Based Cell-Free Toehold Switch Biosensor with a Visual Bioluminescent Output. N. Biotechnol. 2021. https://doi.org/10.1016/j.nbt.2021.09.002.

(22) Compton, J. Nucleic Acid Sequence-Based Amplification. Nature 1991, 354, 56–58.

(23) Lobato, I. M.; O’Sullivan, C. K. Recombinase Polymerase Amplification: Basics, Applications and Recent Advances. TrAC - Trends Anal. Chem. 2018, 98, 19–35. https://doi.org/10.1016/j.trac.2017.10.015.

(24) Piepenburg, O.; Williams, C. H.; Stemple, D. L.; Armes, N. A. DNA Detection Using Recombination Proteins. PLoS Biol. 2006, 4 (7), 1115–1121. https://doi.org/10.1371/journal.pbio.0040204.

(25) Mohsen, M. G.; Kool, E. T. The Discovery of Rolling Circle Amplification and Rolling Circle Transcription. Acc. Chem. Res. 2016, 49 (11), 2540–2550. https://doi.org/10.1021/acs.accounts.6b00417.

(26) Wu, Q.; Suo, C.; Brown, T.; Wang, T.; Teichmann, S. A.; Bassett, A. R. INSIGHT: A Population-Scale COVID-19 Testing Strategy Combining Point-of-Care Diagnosis with Centralized High-Throughput Sequencing. Sci. Adv. 2021, 7 (7). https://doi.org/10.1126/sciadv.abe5054.

(27) Bhadra, S.; Ellington, A. D. A Spinach Molecular Beacon Triggered by Strand Displacement. RNA 2014, 20 (8), 1183–1194. https://doi.org/10.1261/rna.045047.114.

(28) Keightley, M. C.; Sillekens, P.; Schippers, W.; Rinaldo, C.; St. George, K. Real-Time NASBA Detection of SARS-Associated Coronavirus and Comparison with Real-Time Reverse Transcription-PCR. J. Med. Virol. 2005, 77 (4), 602–608. https://doi.org/10.1002/jmv.20498.

(29) Sarcar, S. N.; Miller, D. L. A Specific, Promoter-Independent Activity of T7 RNA Polymerase Suggests a General Model for DNA/RNA Editing in Single Subunit RNA Polymerases. Sci. Rep. 2018, 8 (1), 14–17. https://doi.org/10.1038/s41598-018-32231-6.

(30) Aufdembrink, L. M.; Khan, P.; Gaut, N. J.; Adamala, K. P.; Engelhart, A. E. Highly Specific, Multiplexed Isothermal Pathogen Detection with Fluorescent Aptamer Readout. RNA 2020, 26 (9), 1283–1290. https://doi.org/10.1261/RNA.075192.120.

(31) Biosciences, A. Cell-Free Protein Expression Kit. 2018, 507024 (January), 1–9.

(32) Blum, S. M.; Lee, M. S.; Mgboji, G. E.; Funk, V. L.; Beabout, K.; Harbaugh, S. V.; Roth, P. A.; Liem, A. T.; Miklos, A. E.; Emanuel, P. A.; Walper, S. A.; Chávez, J. L.; Lux, M. W. Impact of Porous Matrices and Concentration by Lyophilization on Cell-Free Expression. ACS Synth. Biol. 2021, 10 (5), 1116–1131. https://doi.org/10.1021/acssynbio.0c00634.

(33) Sun, Z. Z.; Hayes, C. A.; Shin, J.; Caschera, F.; Murray, R. M.; Noireaux, V. Protocols for Implementing an <em>Escherichia Coli</Em> Based TX-TL Cell-Free Expression System for Synthetic Biology. J. Vis. Exp. 2013, No. 79, 3–5. https://doi.org/10.3791/50762.

(34) Ma, D.; Shen, L.; Wu, K.; Diehnelt, C. W.; Green, A. A. Low-Cost Detection of Norovirus Using Paper-Based Cell-Free Systems and Synbody-Based Viral Enrichment. Synth. Biol. 2018, 3 (1), 1–11. https://doi.org/10.1093/synbio/ysy018.

(35) Gibson, D. G.; Young, L.; Chuang, R.-Y.; Venter, J. C.; Hutchison, C. A.; Smith, H. O. Enzymatic Assembly of DNA Molecules up to Several Hundred Kilobases. Nat. Methods 2009, 6 (5), 343–345. https://doi.org/10.1038/nmeth.1318.

(36) Studier, F. W. Protein Production by Auto-Induction in High Density Shaking Cultures. Protein Expr. Purif. 2005, 41 (1), 207–234.

(37) Toparlak, O. D.; Zasso, J.; Bridi, S.; Serra, M. D.; MacChi, P.; Conti, L.; Baudet, M. L.; Mansy, S. S. Artificial Cells Drive Neural Differentiation. Sci. Adv. 2020, 6 (38), 1–13. https://doi.org/10.1126/sciadv.abb4920.

(38) Pardee, K.; Green, A. A.; Takahashi, M. K.; Braff, D.; Lambert, G.; Lee, J. W.; Ferrante, T.; Ma, D.; Donghia, N.; Fan, M.; Daringer, N. M.; Bosch, I.; Dudley, D. M.; O’Connor, D. H.; Gehrke, L.; Collins, J. J. Rapid, Low-Cost Detection of Zika Virus Using Programmable Biomolecular Components. Cell 2016, 165 (5), 1255–1266. https://doi.org/10.1016/j.cell.2016.04.059.

(39) Lavickova, B.; Maerkl, S. J. A Simple, Robust, and Low-Cost Method To Produce the PURE Cell-Free System. ACS Synth. Biol. 2019, 8 (2), 455–462. https://doi.org/10.1021/acssynbio.8b00427.

(40) Grasemann, L.; Lavickova, B.; Elizondo-Cantú, M. C.; Maerkl, S. J. Onepot Pure Cell-Free System. J. Vis. Exp. 2021, 2021 (172), 1–22. https://doi.org/10.3791/62625.

(41) Arce, A.; Guzman Chavez, F.; Gandini, C.; Puig, J.; Matute, T.; Haseloff, J.; Dalchau, N.; Molloy, J.; Pardee, K.; Federici, F. Decentralizing Cell-Free RNA Sensing With the Use of Low-Cost Cell Extracts. Front. Bioeng. Biotechnol. 2021, 9 (August), 1–11. https://doi.org/10.3389/fbioe.2021.727584.

